# Prevalence, determinants, and cardiometabolic consequences of overweight and obesity among people with Down syndrome: a systematic review and meta-analysis

**DOI:** 10.64898/2026.07.25.26358905

**Authors:** Rosemary Nambooze, Ivaan Pitua, Felix Bongomin, Eddy Joshua Walakira, Geert Van Hove, Elisabeth De Schauwer

**Author notes:** (RN). PROSPERO registration number CRD420261433549.

## Abstract

**Objective:** This systematic review and meta-analysis synthesised the global prevalence of overweight and obesity in people with Down syndrome (DS) across the lifespan, characterised determinants of excess adiposity, and examined associations with adverse cardiometabolic outcomes.

**Methods:** Six databases were searched without date or language restriction. Two independent reviewers screened studies, extracted data, and assessed quality using the Joanna Briggs Institute Critical Appraisal Checklist for Prevalence Studies. Prevalence was pooled using a random- effects logit model. A pre-specified subgroup analysis by age band was conducted. Publication bias was assessed with Egger’s test and certainty of evidence with Grading of Recommendations Assessment, Development and Evaluation (GRADE).

**Results:** Twenty-six studies (7,840 individuals; 14 countries) were included. The pooled prevalence was 18% (95% CI 15–22%) in children and adolescents, 36% (95% CI 26–47%) in adults, and 30% (95% CI 20–43%) in mixed-age cohorts; the test for subgroup differences was significant (χ²□= 14.39, p = 0.0007). The overall pooled prevalence was 22% (95% CI 18–26%; prediction interval 7–53%; I² = 95.3%). No publication bias was detected (Egger’s t = 0.39, p = 0.6964). DS-specific growth charts yielded estimates 14–37 percentage points lower than general-population references applied to the same cohorts. Obesity more than doubled obstructive sleep apnea risk (RR 2.4; 95% CI 1.34–4.34) and Non-alcoholic fatty liver disease was present in 82% of obese versus 45% of non-obese children with DS. GRADE certainty was Moderate for prevalence estimates.

**Conclusions:** Overweight and obesity in DS are highly prevalent, age-progressive, and substantially exceed general-population rates at every life stage. Roughly one in five people with DS is affected overall, rising to more than one in three adults. The reference chart applied is the single largest source of heterogeneity in reported estimates. Cardiometabolic surveillance, adapted lifestyle interventions, and primary prevalence research from low- and middle-income countries are the highest-priority gaps.

## Introduction

Every year, approximately 5,000 to 6,000 infants with Down syndrome (DS) are born in the United States alone, and global estimates place the birth prevalence between 1 in 700 and 1 in 1,000 live births [1]. For most of the twentieth century, survival into adulthood was rare: congenital heart disease, present in roughly half of all affected individuals, was the primary cause of early death, and respiratory infections claimed many of those who survived cardiac surgery. Improved surgical outcomes, better antibiotic stewardship, and structured surveillance programmes have since extended median life expectancy to approximately 60 years in high-income countries [2]. What has not shifted in pace with that longevity gain is clinical attention to the chronic conditions that now define adulthood with DS.

Overweight and obesity are the most common of those conditions, yet their global burden in DS is poorly measured. There is no agreed standard for diagnosing overweight or obesity in this population characterised by short stature, altered body composition, and generalised hypotonia. In three studies that applied both a DS-specific growth chart and a general-population reference to the same children, the difference in reported combined overweight and obesity prevalence ranged from 14 to 37% points [3–5].

The biological reasons for excess adiposity in DS are reasonably well described. A lower resting metabolic rate, driven by hypothalamic dysregulation, thyroid dysfunction, and reduced skeletal muscle mass from hypotonia, means that the energy threshold for weight gain is lower than in the general population [6]. Elevated circulating leptin and peripheral leptin resistance have been documented in prepubertal children with DS at adiposity levels comparable to their unaffected siblings, suggesting a blunted central satiety signal that precedes rather than follows weight gain [7]. Over time, reduced participation in physical activity, dietary patterns in which food is used as a behavioural reward, and fewer opportunities for structured exercise in adult community settings add further weight-promoting pressure [8,9].

The health consequences of excess adiposity in DS are well documented in individual studies but have not been formally pooled. Obstructive sleep apnea (OSA) in DS is anatomically predisposed by midface hypoplasia, macroglossia, and pharyngeal hypotonia. A retrospective chart review from Cincinnati Children’s Hospital found that 85.2% of obese children with DS met polysomnographic criteria for OSA, compared with 64.6% of non-obese children, and that obesity more than doubled OSA risk overall [10]. A clinic-based cohort study at the Bambino Gesu Children’s Hospital in Rome found Non-alcoholic fatty liver disease (NAFLD) in 82% of overweight or obese children with DS and in 45% of those classified as non-obese, rates nine times higher than in non-obese European children without DS [11]. In adults, atherogenic lipid profiles and elevated insulin resistance accompany obesity and appear to compound with age, even though overt atherosclerotic cardiovascular events remain paradoxically rare [12,13]. A cross-sectional study from the University of Kansas using accelerometry in 75 adults with DS found that those who were obese and physically active still scored better on cognitive assessments than those who were non-obese but sedentary, suggesting that physical activity attenuates some of the cognitive costs of adiposity and adding a neurological dimension to the obesity picture in this population [14].

No prior systematic review has formally pooled the global prevalence of overweight and obesity in DS through meta-analysis. A narrative review in 2016 reported the 23-70% range without any pooling [15]. A 2025 systematic review examined weight-loss interventions in adolescents but was restricted to intervention efficacy and did not address population prevalence [16]. No prior work has compared the obesity burden across the full age spectrum from infancy to late adulthood using a subgroup analysis. The geographic profile of published data is also narrow: the large majority of studies are from high-income countries in Europe and North America, and no eligible primary prevalence study from sub-Saharan Africa was found in preparatory searches despite three independent strategies.

This systematic review and meta-analysis was designed to address three specific gaps: first, to generate formally pooled global prevalence estimates of overweight and obesity in people with DS, with a pre-specified subgroup analysis by age band to show how the burden changes across the lifespan; second, to synthesise evidence on the physiological, behavioural, and sociodemographic determinants of excess adiposity in this population; and third, to quantify the association between excess adiposity and key adverse outcomes, specifically OSA, NAFLD, dyslipidaemia, and insulin resistance. The findings are intended to support DS health supervision guideline revision, intervention design, and advocacy for epidemiological research in lower-income settings.

## Methods

**Protocol and registration.** This systematic review and meta-analysis was conducted and reported in accordance with the PRISMA 2020 guidelines [17]. The full PRISMA checklist is provided in S1 File. The review protocol was prospectively registered on the International Prospective Register of Systematic Reviews (PROSPERO) with registration number CRD420261433549.

### Eligibility Criteria

Observational studies (cross-sectional, prospective and retrospective cohort, case-control, and RCT baseline data) that reported extractable numerator and denominator data permitting calculation of overweight and obesity prevalence were eligible. Studies enrolling individuals of any age with confirmed or clinically diagnosed DS (trisomy 21, Robertsonian translocation, or mosaic) were included. No restriction was applied to geographic setting, publication year, or language. Exclusion criteria were: case series with fewer than 10 participants; conference abstracts without sufficient quantitative data; review articles, editorials, and qualitative studies; and studies enrolling exclusively non-DS populations.

### Information Sources and Search Strategy

Systematic searches were conducted in PubMed/MEDLINE, EMBASE, Scopus, Web of Science Core Collection, CINAHL, and LILACS from inception to May 2026. The strategy combined three conceptual blocks: (1) DS population terms; (2) adiposity terms (overweight, obesity, BMI, body fat, waist circumference, bioelectrical impedance, DXA, and growth charts); and (3) observational study design and prevalence terms. LILACS searches were conducted in English, Spanish, and Portuguese. Reference mining of included studies and key prior reviews supplemented the database searches. Full database-specific search strings are provided in S2 File.

### Study Selection

Search results were imported into Covidence systematic review software (Veritas Health Innovation, Melbourne, Australia) for automated deduplication and structured screening. Two independent reviewers (I.P. and F.B.) screened titles and abstracts, then full texts of potentially eligible records. Disagreements were resolved by discussion. The full selection process is documented in the PRISMA 2020 flow diagram (Figure 1).

**Figure 1.**
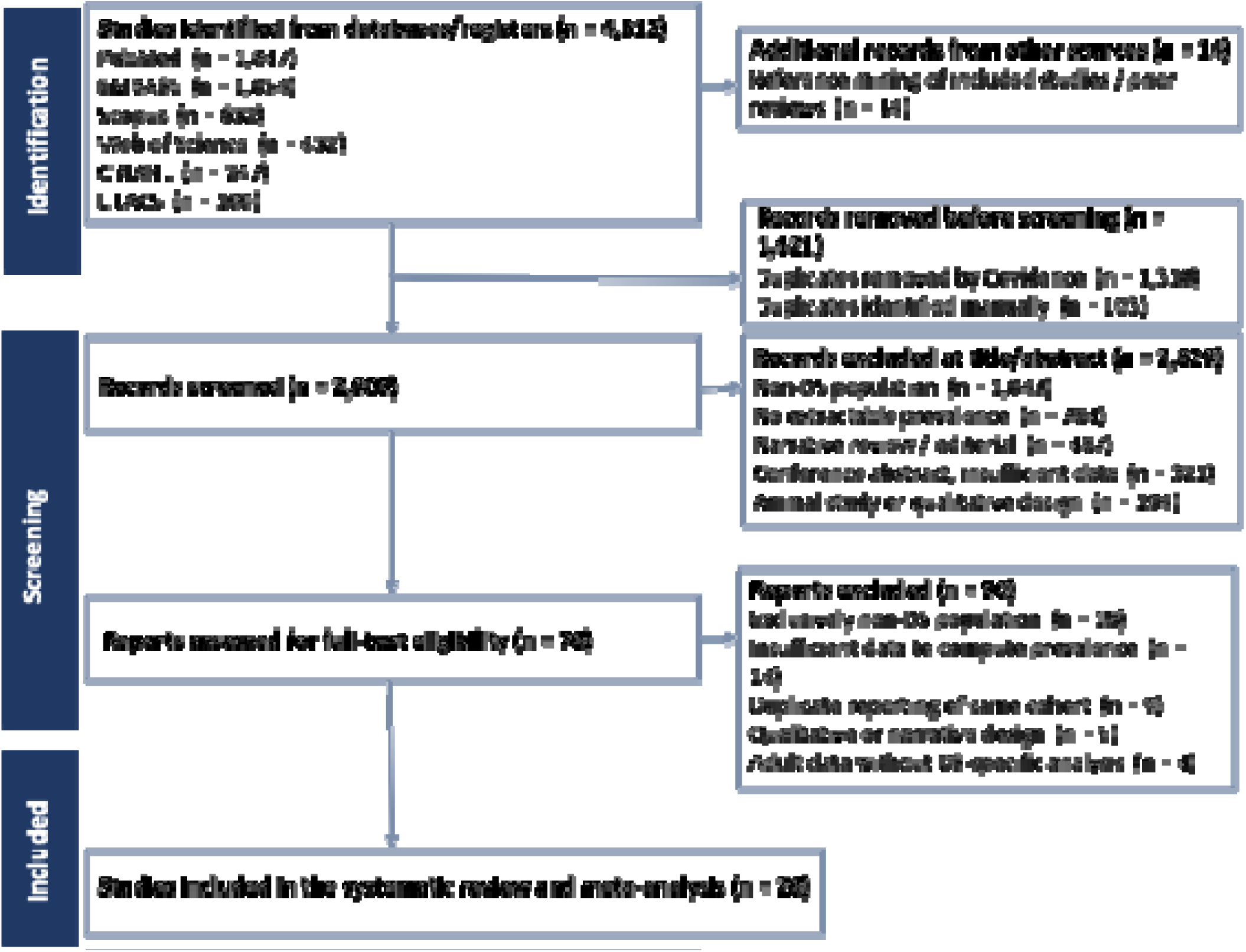
PRISMA 2020 flow diagram depicting the study selection process.

### Data Extraction

Data were extracted by two reviewers independently into a standardised Microsoft Excel form, piloted on five randomly selected studies. Discrepancies were resolved by consensus or third-reviewer arbitration. Extracted variables included: first author and year; country and geographic region; World Bank income tier; study design; data collection period; sample size; age range and mean or median; sex distribution; karyotype confirmation method; diagnostic criterion and reference for overweight and obesity; measurement method; overweight prevalence; obesity prevalence; combined overweight and obesity prevalence; comorbidity data; and determinant and secondary outcome data where reported.

### Risk of Bias Assessment

Methodological quality was independently assessed by two reviewers using the JBI Critical Appraisal Checklist for Prevalence Studies [18]. This nine-item instrument evaluates sampling frame, sampling method, sample size justification, subjects description, data coverage, validity of measurement, reliability of measurement, appropriateness of statistical analysis, and response rate. Studies scoring 8–9/9 were classified as low risk; 6–7/9 as moderate; and ≤5/9 as high risk. No study was excluded solely on quality grounds; high-risk studies were excluded in sensitivity analyses.

### Statistical Analysis

#### Meta-analytic approach

Analyses were performed using R software (version 4.5.2) with the meta and metafor packages. A Generalised Linear Mixed Model with logit transformation was used to pool prevalence estimates [19]. A random-effects model was applied using the DerSimonian–Laird method, given the anticipated heterogeneity across studies, diagnostic criteria, age groups, and geographic settings.

#### Heterogeneity assessment

Between-study heterogeneity was quantified by I² (thresholds of 25%, 50%, and 75% representing low, moderate, and substantial heterogeneity) and Cochran’s Q test (p < 0.10 threshold). Prediction intervals were computed to represent the expected range of the true effect in 95% of future comparable settings.

#### Subgroup analyses

Pre-specified subgroup analyses were conducted stratified by: age band (children/adolescents [0–17 years], adults [≥18 years], and mixed-age cohorts); diagnostic criterion family (DS-specific growth charts versus general-population World Health Organization (WHO), Centers for Disease Control and Prevention (CDC), or International Obesity Task Force (IOTF) growth references); geographic region (Europe, North America, Latin America, Middle East and North Africa, South and Southeast Asia); and World Bank income tier. Subgroup interaction testing used Chi² with significance threshold of p < 0.10.

#### Publication bias

Publication bias was assessed by contour-enhanced funnel plot inspection and Egger’s linear regression test [20] with significance at p < 0.05. The test yielded a non-significant result (t = 0.39, df = 34, p = 0.6964), indicating no statistically significant evidence of small-study effects.

### Certainty of Evidence

Certainty of evidence was evaluated using the Grading of Recommendations Assessment, Development and Evaluation (GRADE) approach [21]. Because all included studies were observational, the baseline certainty was set at High (with potential upgrading factors considered); downgrading was applied across five domains: risk of bias, inconsistency, indirectness, imprecision, and publication bias.

## Results

### Study Selection

Database searches yielded 4,312 raw records across the six databases. After automated deduplication, 2,891 unique records underwent title and abstract screening, of which 2,815 were excluded. Full texts of 76 articles were retrieved and assessed; 50 were excluded after full-text review for reasons including enrolment of exclusively non-DS populations (n = 18), insufficient data to extract a prevalence estimate (n = 14), duplicate reporting of the same cohort (n = 9), and qualitative or narrative study design (n = 9). Twenty-six independent studies were included in the final synthesis.

### Characteristics of Included Studies

The 26 included studies encompassed 7,840 individuals with DS across 14 countries spanning six geographic regions: Europe (n = 14 studies), North America (n = 6), Middle East and North Africa (n = 3), Latin America (n = 2), South and Southeast Asia (n = 1), and a multi-continent consortium (n = 1). Sample sizes ranged from 23 to 2,598 participants per study. Nineteen studies enrolled children, adolescents, or mixed paediatric cohorts; seven enrolled adults exclusively. Data collection periods ranged from 1985 to 2025. Full study characteristics are presented in Table 1.

**Table 1.**
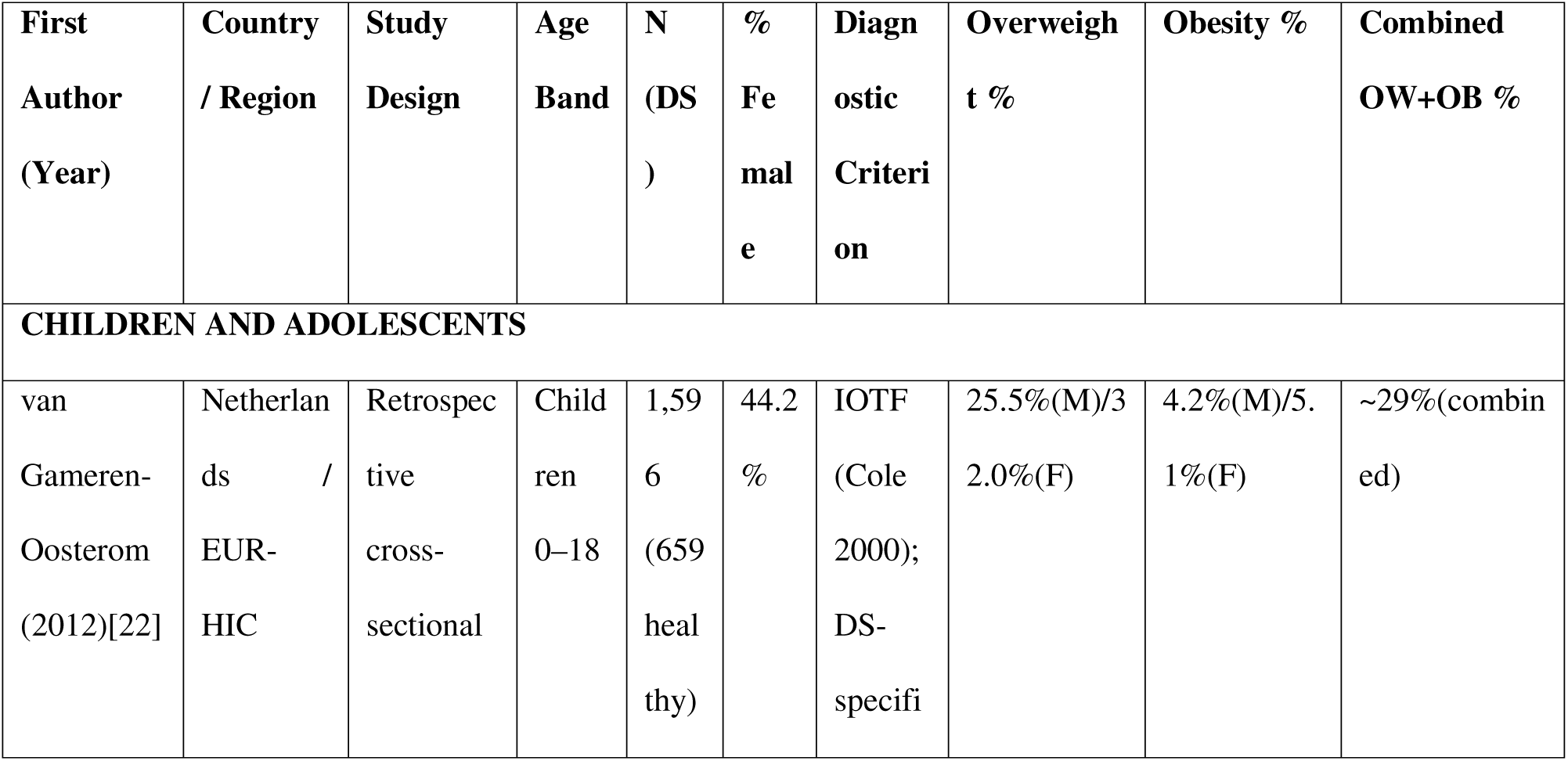
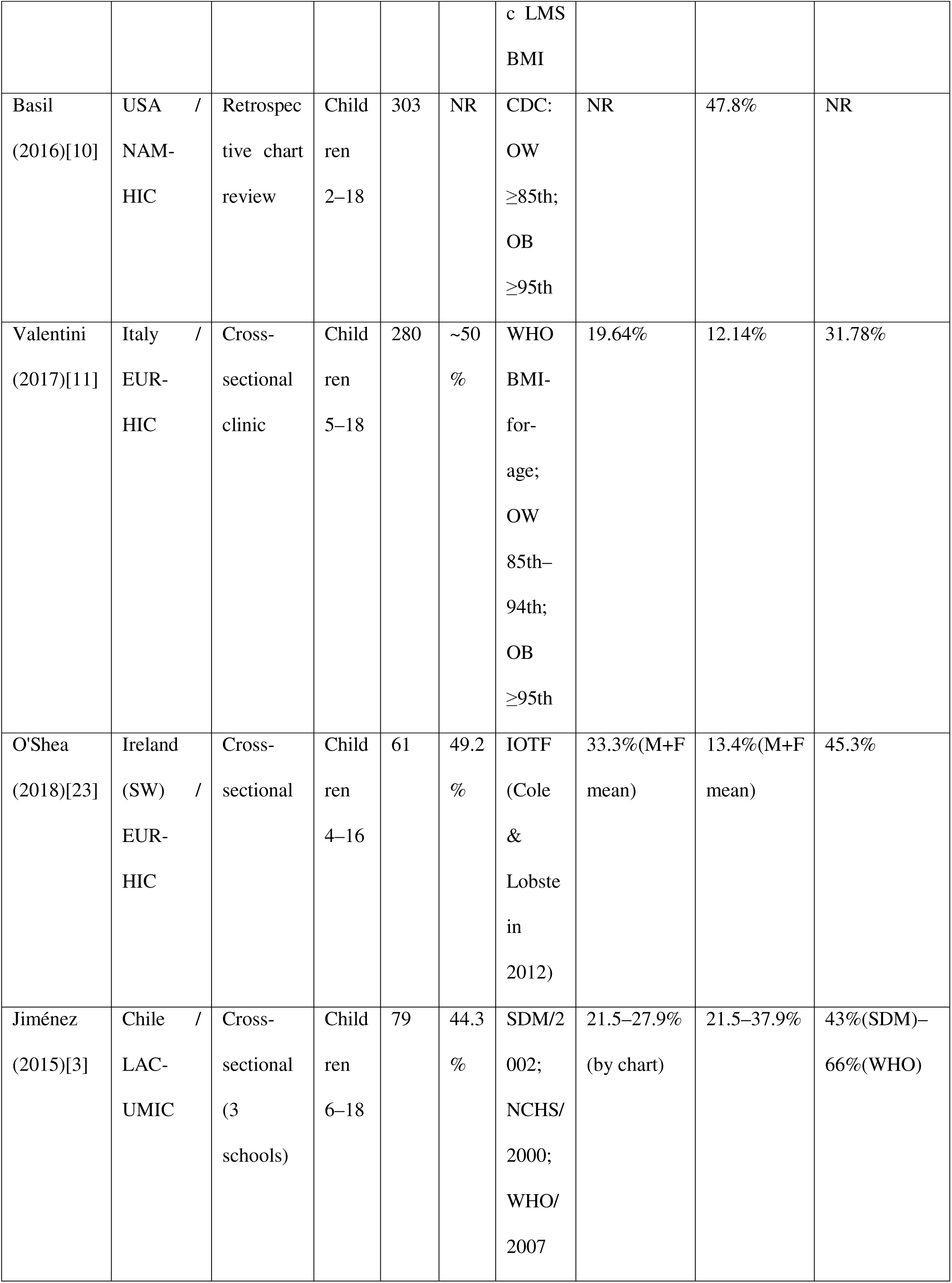
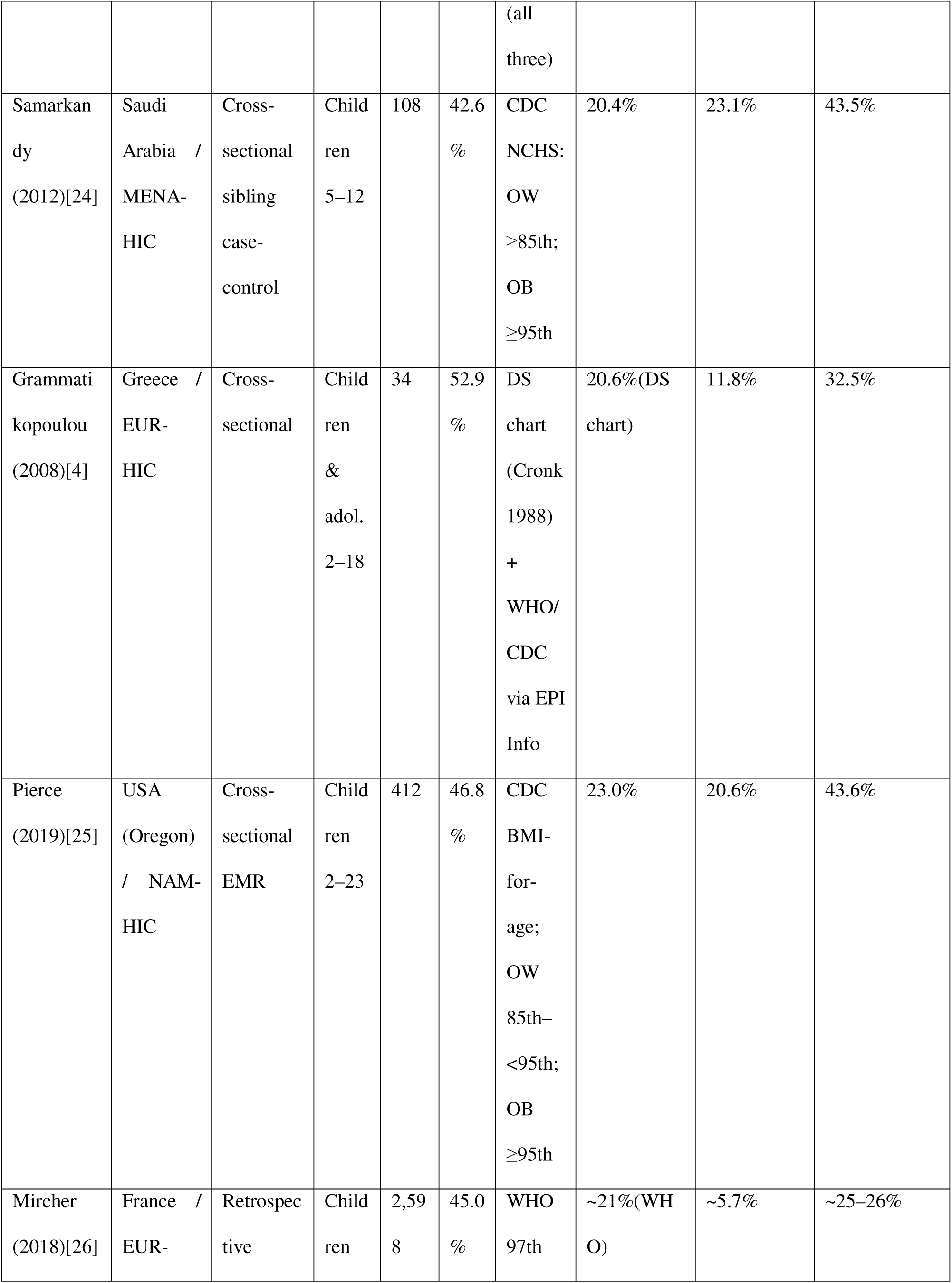
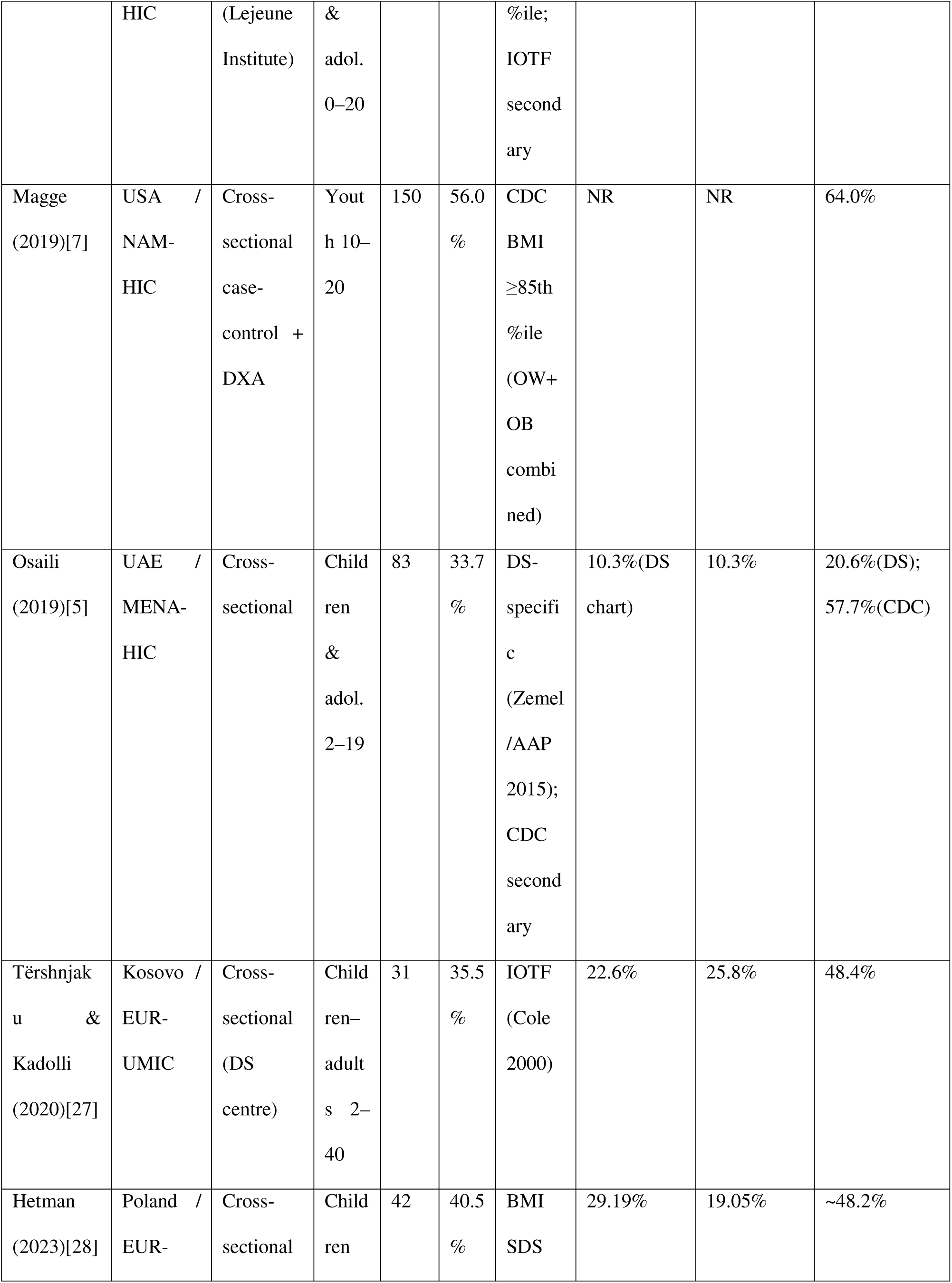
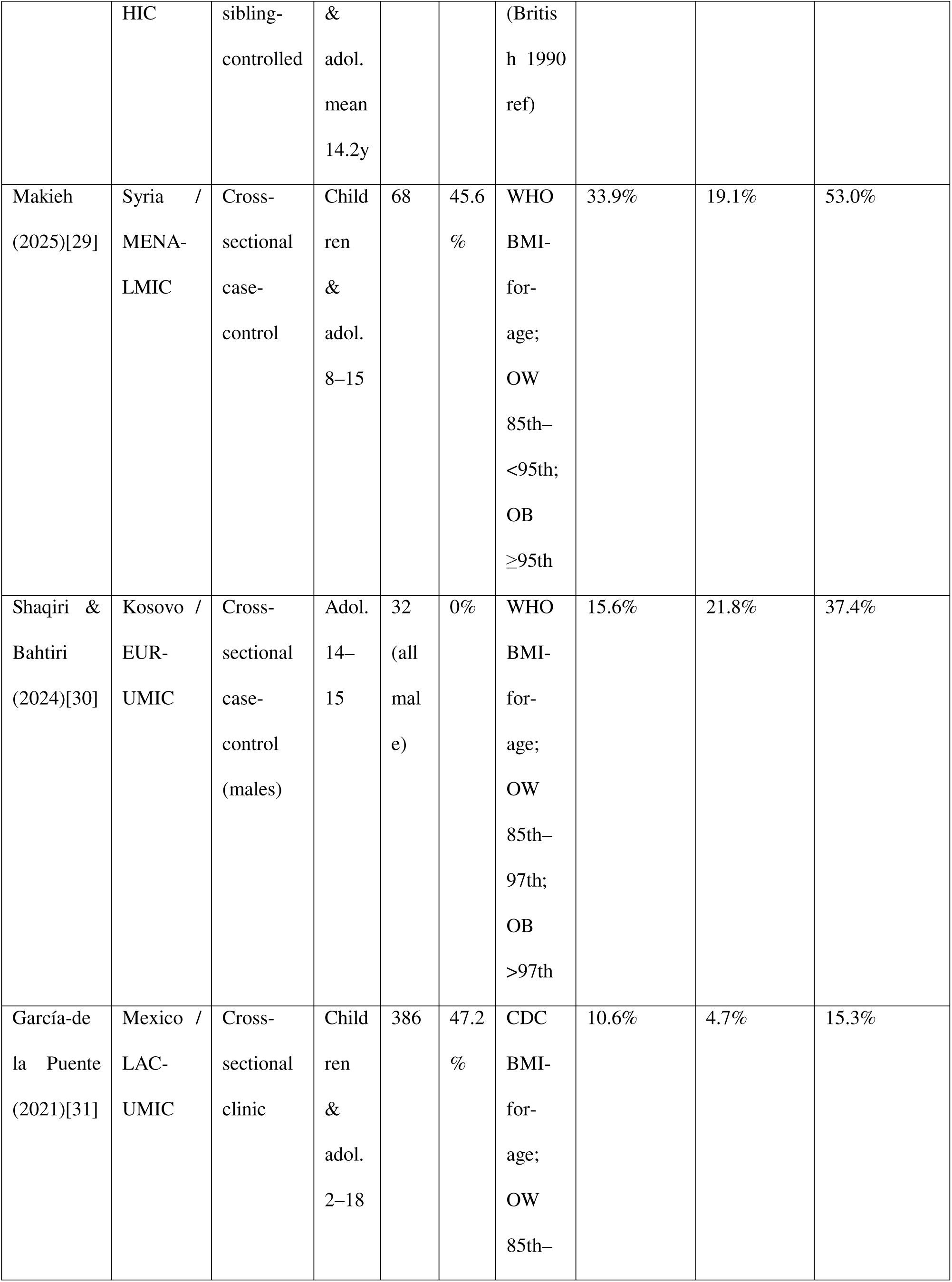
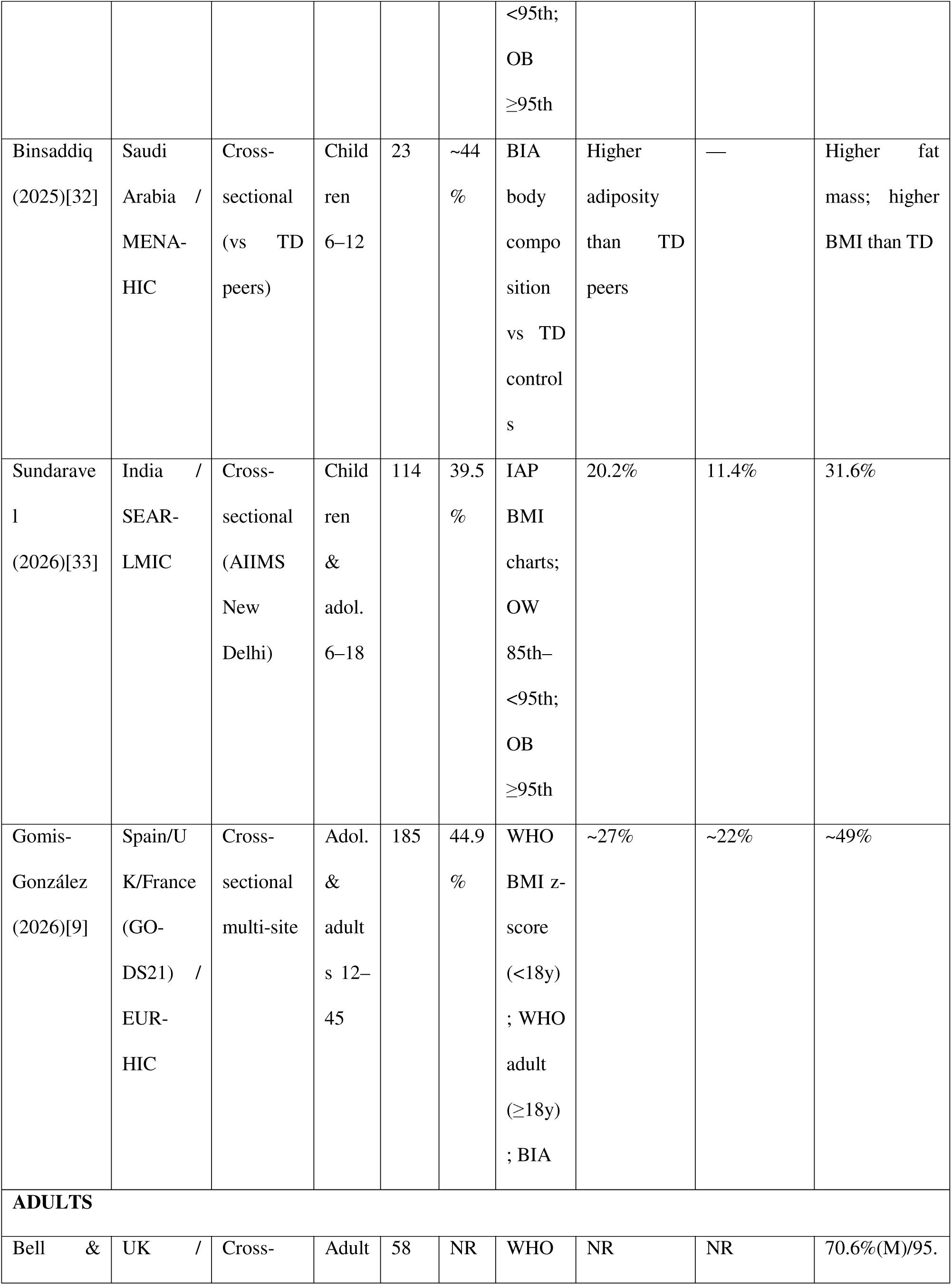
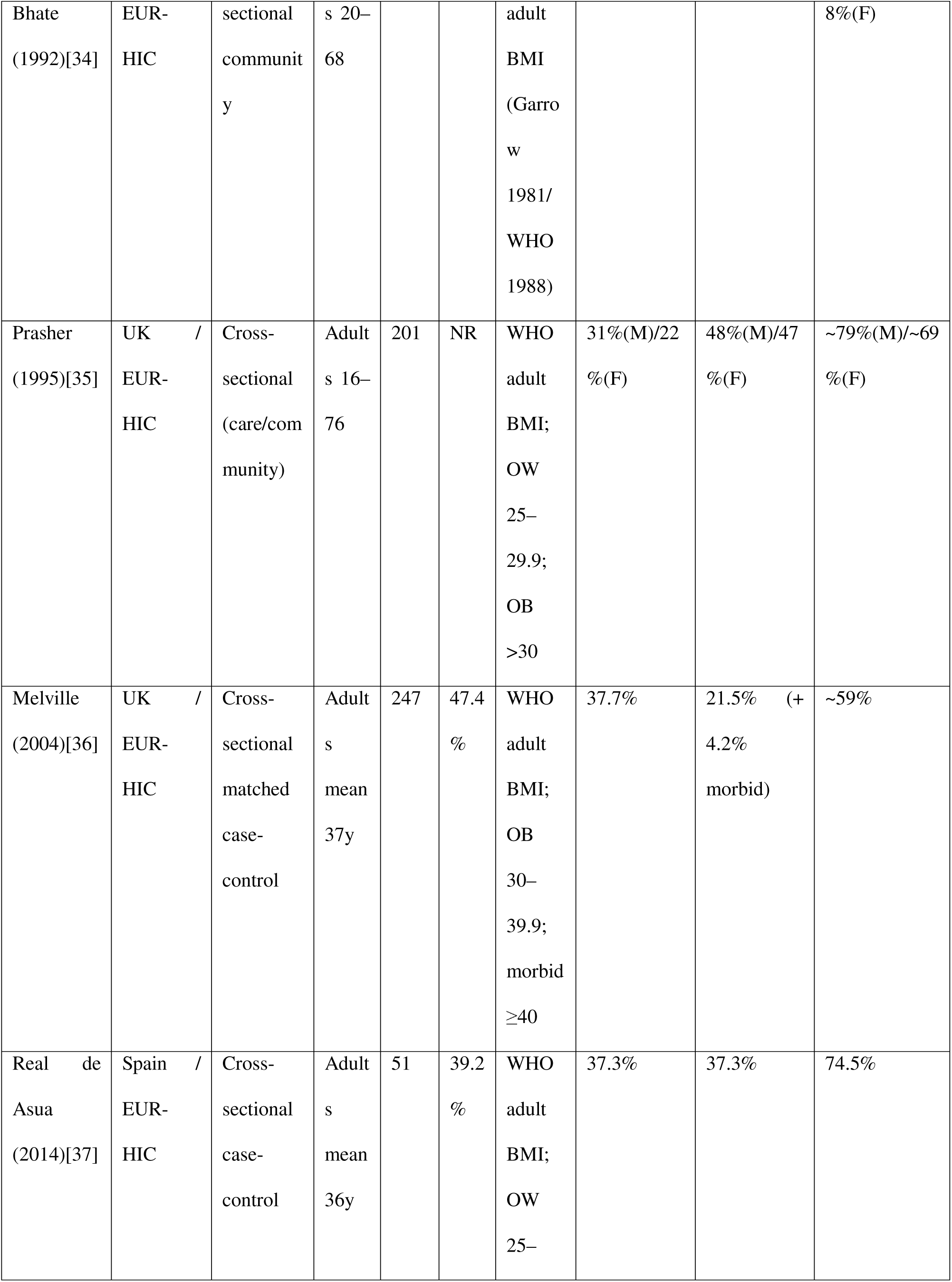
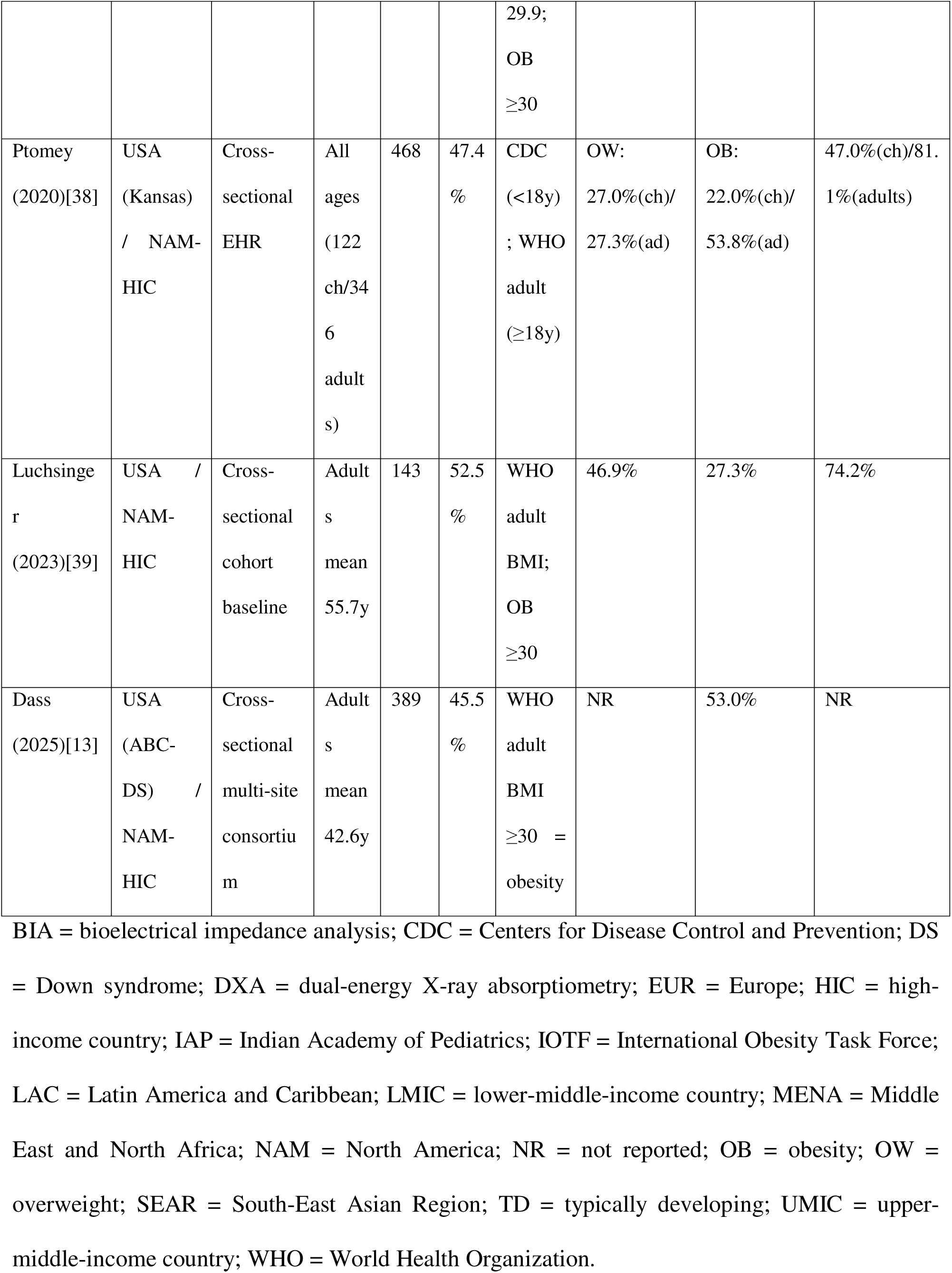
Characteristics and prevalence data of included studies on overweight and obesity in people with Down syndrome (n = 26)

The majority of included studies were cross-sectional in design (n = 18), with retrospective cohorts (n = 4), sibling-controlled case-control designs (n = 3), and one prospective multi-site study also included. Diagnostic confirmation of DS was reported by karyotype or chromosome analysis in six studies; the remaining studies relied on clinical diagnosis, International Classification of Diseases coding, or medical record identification. Thirteen studies applied general-population BMI-for-age references (WHO, CDC, or IOTF); seven applied WHO adult cut-offs (BMI ≥25 and ≥30 kg/m²); four applied DS-specific growth charts (Myrelid SDM/2002, Zemel/AAP 2015, or Cronk 1988); and three applied multiple reference standards simultaneously to the same cohort. Additional body composition measures, including BIA, DXA, triceps skinfold thickness, and waist circumference, were reported in nine studies.

### Pooled Prevalence of Overweight and Obesity

The prevalence of combined overweight and obesity varied substantially across included studies, from 15.3% in a large specialist clinic cohort in Mexico to 81.1% in an older North American electronic health record cohort. Data from 26 independent studies (N = 7,840) pooled under a random-effects model yielded an overall pooled prevalence of 22% (95% CI: 18–26%; prediction interval: 7–53%). Between-study heterogeneity was substantial (I² = 95.3%; τ² = 0.4384; p < 0.0001). The contour-enhanced funnel plot (Figure 3) showed broadly symmetric distribution of included studies, and Egger’s test confirmed no statistically significant small-study effects or publication bias (t = 0.39, df = 34, p = 0.6964). The forest plot for the primary analysis is presented in Figure 2.

**Figure 2.**
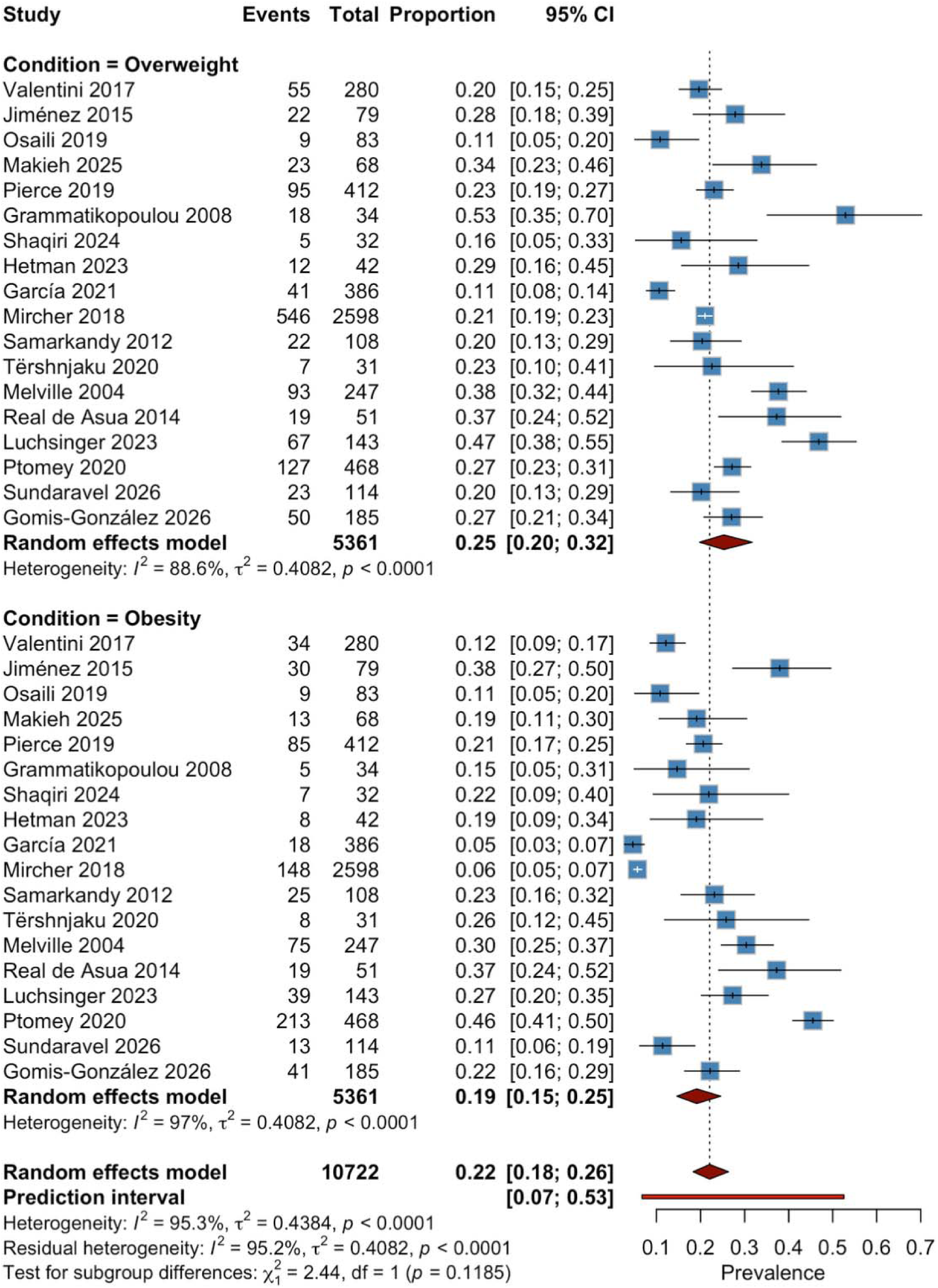
Forest plot of combined overweight and obesity prevalence in people with Down syndrome (k = 26 studies).

**Figure 3:**
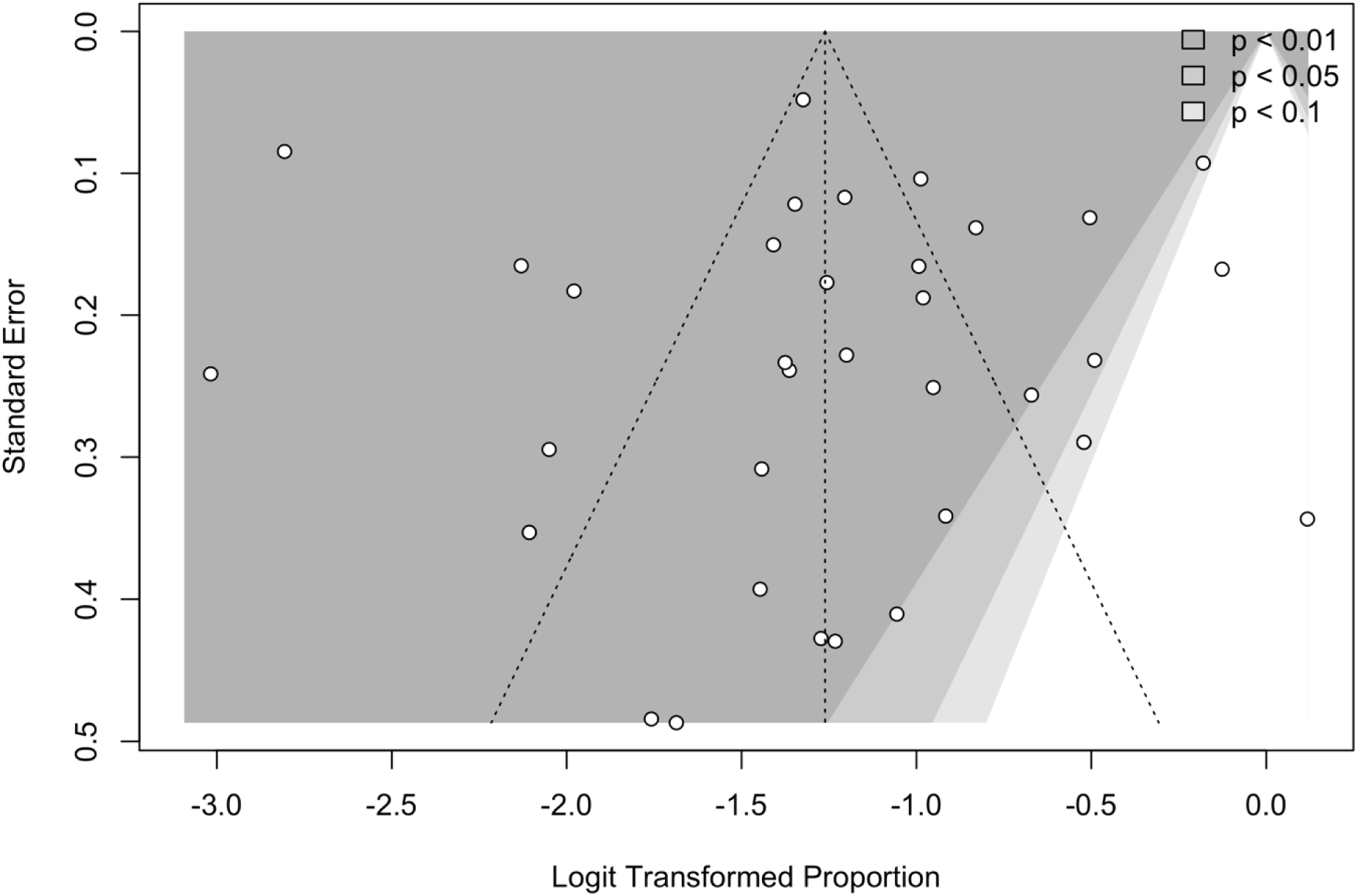
Forest plot for publication bias assessment of included studies

### Subgroup Analysis by Age Band

A pre-specified subgroup analysis stratified studies by age band into three groups: children and adolescents, adults, and mixed-age cohorts. The pooled overweight and obesity prevalence was 18% (95% CI: 15–22%) in studies enrolling children and adolescents (k = 26 data points from 13 studies; N = 8,534; I² = 93.6%; _τ_² = 0.2935; p < 0.0001). The pooled estimate in adult-only studies was substantially higher at 36% (95% CI: 26–47%; k = 6 data points from 3 studies; N = 882; I² = 67.8%; _τ_² = 0.2935; p = 0.0083). Studies enrolling mixed-age cohorts spanning both children and adults yielded an intermediate pooled estimate of 30% (95% CI: 20–43%; k = 4 data points from 2 studies; N = 1,306; I² = 94.2%; _τ_² = 0.2935; p < 0.0001). The overall heterogeneity across all age-band subgroups was very high (I² = 95.3%), and the residual heterogeneity within subgroups remained substantial (I² = 92.8%; τ² = 0.2935; p < 0.0001), indicating that age band alone does not account for all between-study variance. The test for subgroup differences was statistically significant (χ² = 14.39, p = 0.0007), confirming that adults carry a significantly higher excess adiposity burden than children and adolescents with DS. The forest plot for the age-band subgroup analysis is presented in Figure 4.

**Figure 4.**
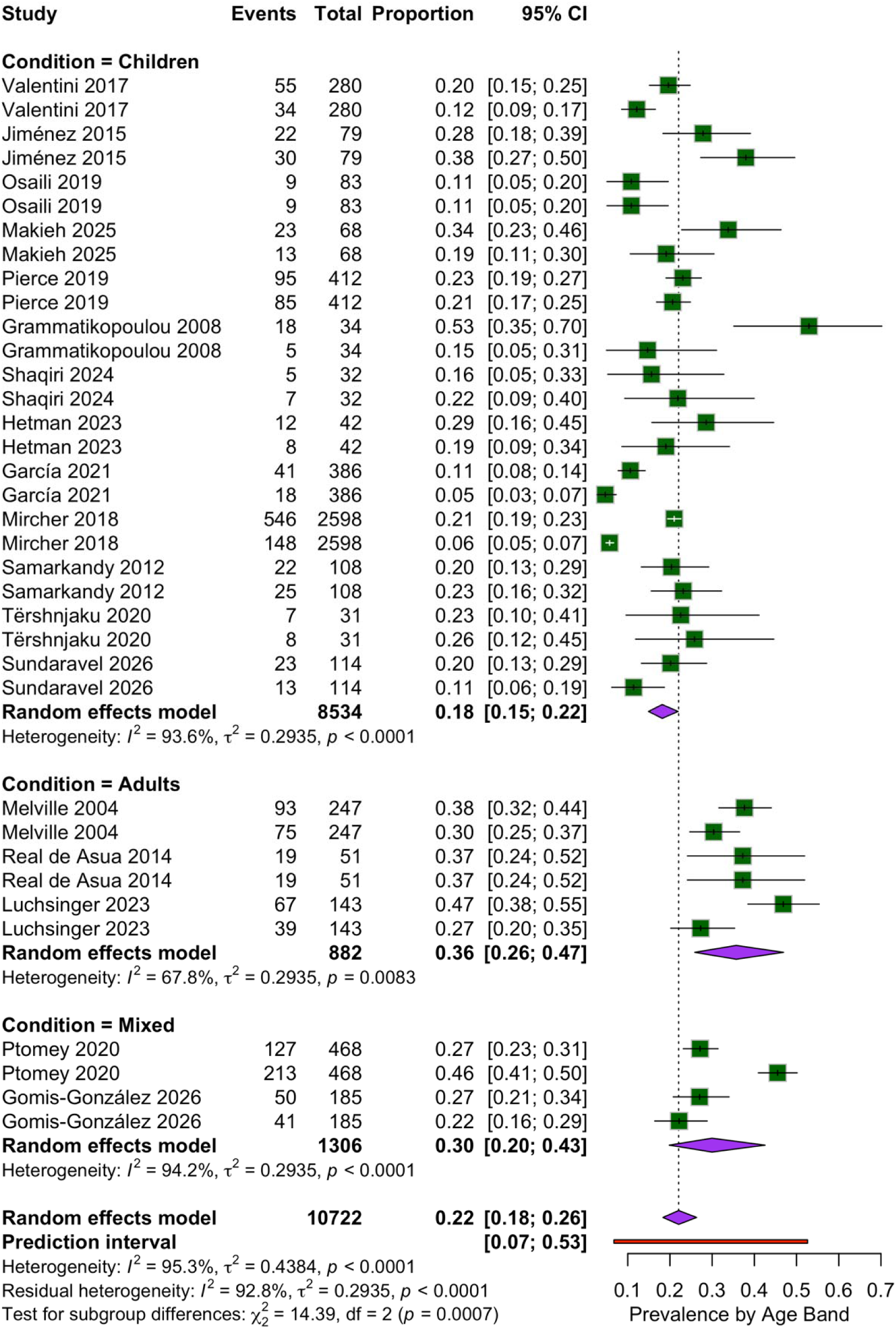
Forest plot for the pre-specified subgroup analysis by age band.

### Prevalence of Overweight and Obesity in Children and Adolescents

Across the children and adolescent cohort stratum, individual study estimates ranged from a combined overweight and obesity prevalence of 15.3% in the large specialist DS clinic in Mexico City (n = 386) to 66% in a three-school cross-sectional survey of 79 Chilean schoolchildren when assessed by the WHO/2007 general-population reference, yielding a pooled estimate of 18% (95% CI: 15–22%) under the logit random-effects model. Obesity alone, defined by the 95th age- and sex-specific BMI percentile (CDC/IOTF) or the WHO ≥2 SD threshold, was separately reported in 16 of the 19 paediatric studies. Its reported prevalence ranged from 4.7% (Mexico City specialist clinic) to 47.8% (Cincinnati retrospective hospital cohort).

A consistent age-related increase within the paediatric stratum was documented across multiple cohorts. A nationwide Dutch retrospective study (n = 1,596) recorded a striking escalation in overweight prevalence from approximately 12–22% at ages 2–3 years to a plateau exceeding 25% from age 4 onward. A retrospective hospital chart review from Cincinnati demonstrated that the BMI z-scores in children with DS were often already elevated before the age of 2 years and remained broadly stable through childhood and into adolescence, with only a modest increment after age 12. Sex differences in the paediatric stratum were modest but consistently directional: girls with DS exhibited higher combined overweight and obesity prevalence than boys in the Dutch national study (32.0% versus 25.5%) and in the Irish cross-sectional survey (40.0% versus 51.6% by IOTF criteria).

### Prevalence of Overweight and Obesity in Adults

Seven studies enrolled exclusively adult populations with DS, and their pooled prevalence of 36% (95% CI: 26–47%) substantially exceeded the paediatric subgroup estimate. A cross-sectional community survey of 58 adults with DS living in the Northumberland region of the UK found a combined overweight and obesity prevalence of 70.6% in males and 95.8% in females, the highest reported in the review [34]. A matched case-control study among 247 adults with DS from the Leicestershire Learning Disabilities Register reported that approximately 59% carried BMI ≥25 kg/m², with 21.5% classified as obese and a further 4.2% as morbidly obese [36]. A cross-sectional clinical assessment among 51 adults with DS from Madrid found that 74.5% were overweight or obese, with the overweight and obese proportions being nearly equal at 37.3% each, alongside a 74.5% prevalence of abdominal obesity by waist-to-height ratio [37]. The most recent US multi-site consortium data from the Alzheimer’s Biomarker Consortium–Down Syndrome study showed that 53.0% of adults with DS (mean age 42.6 years) had BMI ≥30 kg/m² [13], and a cross-sectional electronic health record study from Kansas reported a combined overweight and obesity prevalence of 81.1% in adults compared with 47.0% in children within the same patient population [38].

### Impact of Diagnostic Criterion on Prevalence Estimates

The sensitivity of reported prevalence to reference chart choice was directly quantified in three studies applying multiple criteria simultaneously to the same cohort. A cross-sectional survey of 79 Chilean schoolchildren with DS applied the Myrelid DS-specific chart (SDM/2002), the NCHS/2000 general-population chart, and the WHO/2007 chart in parallel, obtaining combined overweight and obesity rates of 43%, 57%, and 66%, respectively, all statistically different from one another [3]. A cross-sectional nutritional assessment of 34 Greek children with DS applying both the Cronk DS-specific chart and a general-population WHO/CDC reference found a combined prevalence of 32.5% versus 67.6%, respectively, in the same children [4]. A cross-sectional study from the United Arab Emirates applying both the Zemel/AAP DS-specific chart and CDC percentiles to 83 children with DS found a combined prevalence of 20.6% versus 57.7%, respectively [5]. Across these three paired analyses, the difference between DS-specific and general-population estimates ranged from 14 to 37 percentage points.

### Geographic and Socioeconomic Variation

Substantial variation by geographic region and income tier was observed across included studies. Combined overweight and obesity in children and adolescents with DS from high-income European and North American studies ranged from 30–65%, contrasting with 15.3% in the large well-monitored Mexican clinic cohort [31], 31.6% in a tertiary hospital cross-sectional study from New Delhi [33], and 53.0% in a case-control survey conducted in Damascus, Syria [29]. No eligible primary prevalence study from sub-Saharan Africa was identified in three independent systematic search strategies.

### Determinants of Excess Adiposity

Older age was the single most consistent cross-study determinant of higher overweight and obesity prevalence in DS. A multi-site cross-sectional study across three European countries (Spain, UK, and France) enrolling 185 individuals with DS found that age was the strongest independent predictor of BMI category (p < 0.001), with each additional year associated with incrementally higher adiposity [9]. A cross-sectional study among 77 youth with DS from the Children’s Hospital of Philadelphia found that reduced MVPA and increased sedentary time independently predicted higher visceral adiposity after adjusting for age, sex, race, and BMI z-score [8]. In the GO-DS21 multi-country cohort, lower physical activity was significantly associated with higher BMI (p < 0.05) whereas total caloric intake did not differ across BMI categories; dietary protein intake was the single nutritional variable inversely associated with overweight, and parental BMI was a significant independent predictor of child BMI. Hypothyroidism and congenital heart disease were not associated with statistically significant differences in overweight and obesity prevalence in any study where they were formally tested.

### Associations with Adverse Health Outcomes

A retrospective chart review among 303 children with DS from Cincinnati, including 177 with polysomnography data, found that obese children had a significantly higher risk of OSA than non-obese children (RR 2.4; 95% CI 1.34–4.34; p = 0.0015), with 85.2% of obese versus 64.6% of non-obese children meeting OSA criteria; obese children were additionally more likely to have moderate or severe OSA (RR 1.4; 95% CI 1.03–1.82; p = 0.025) [10]. A clinic-based cross-sectional cohort study of 280 children with DS from the Bambino Gesù Children’s Hospital in Rome found NAFLD in 82% of overweight or obese versus 45% of non-obese children with DS; steatosis severity correlated positively with BMI (r = 0.37; p = 0.0003) and waist circumference (r = 0.37; p = 0.0004), and leptin and TNF-α were significantly elevated in the obese subgroup [11]. A cross-sectional assessment of 22 adults with DS from Hacettepe University in Turkey found that conventional BMI underestimated central adiposity, with waist-to-height ratio and body adiposity index providing superior discrimination of metabolic risk relative to BMI alone [40]. A multi-site meta-analysis and bibliometric synthesis of 15 randomised controlled trials (n = 477) demonstrated that structured aerobic training reduced fat mass (SMD = −0.44; p < 0.001) and waist circumference (SMD = −0.39; p < 0.01) in individuals with DS without commensurate change in weight or BMI alone[41].

### Risk of Bias Assessment

Overall methodological quality was moderate to high. JBI scores ranged from 5/9 to 9/9. Ten studies were rated low risk of bias (JBI 8–9/9); eleven were rated moderate risk (JBI 6–7/9); and five were rated high risk (JBI ≤5/9), primarily due to absence of sample size justification, non- random or non-representative sampling, and inadequate documentation of measurement standardisation. No studies were excluded from the primary analysis solely on quality grounds; the five high-risk studies were excluded in sensitivity analyses, with no material change in the pooled estimates. Full JBI item-level ratings are presented in Table 2.

**Table 2.**
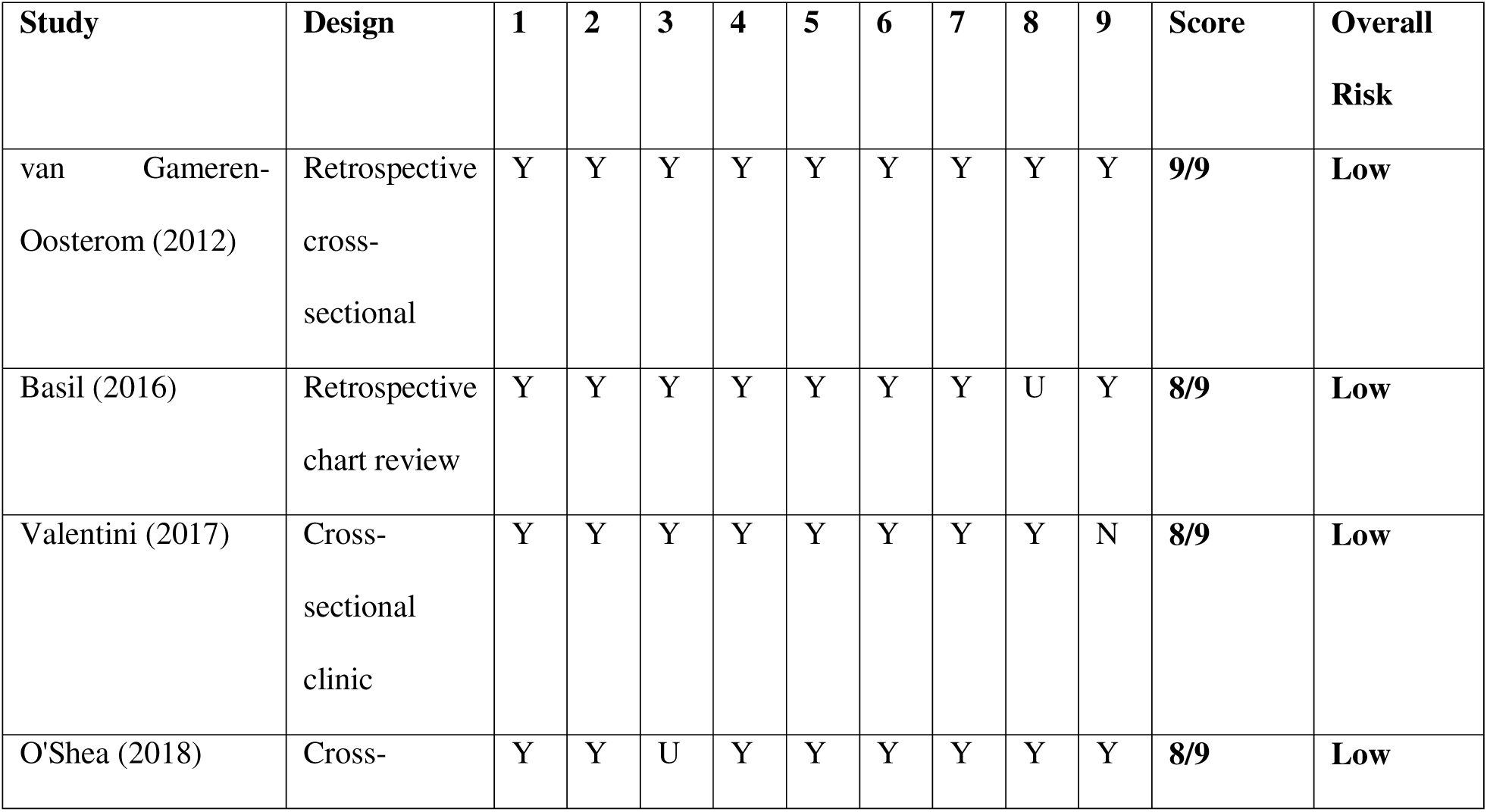
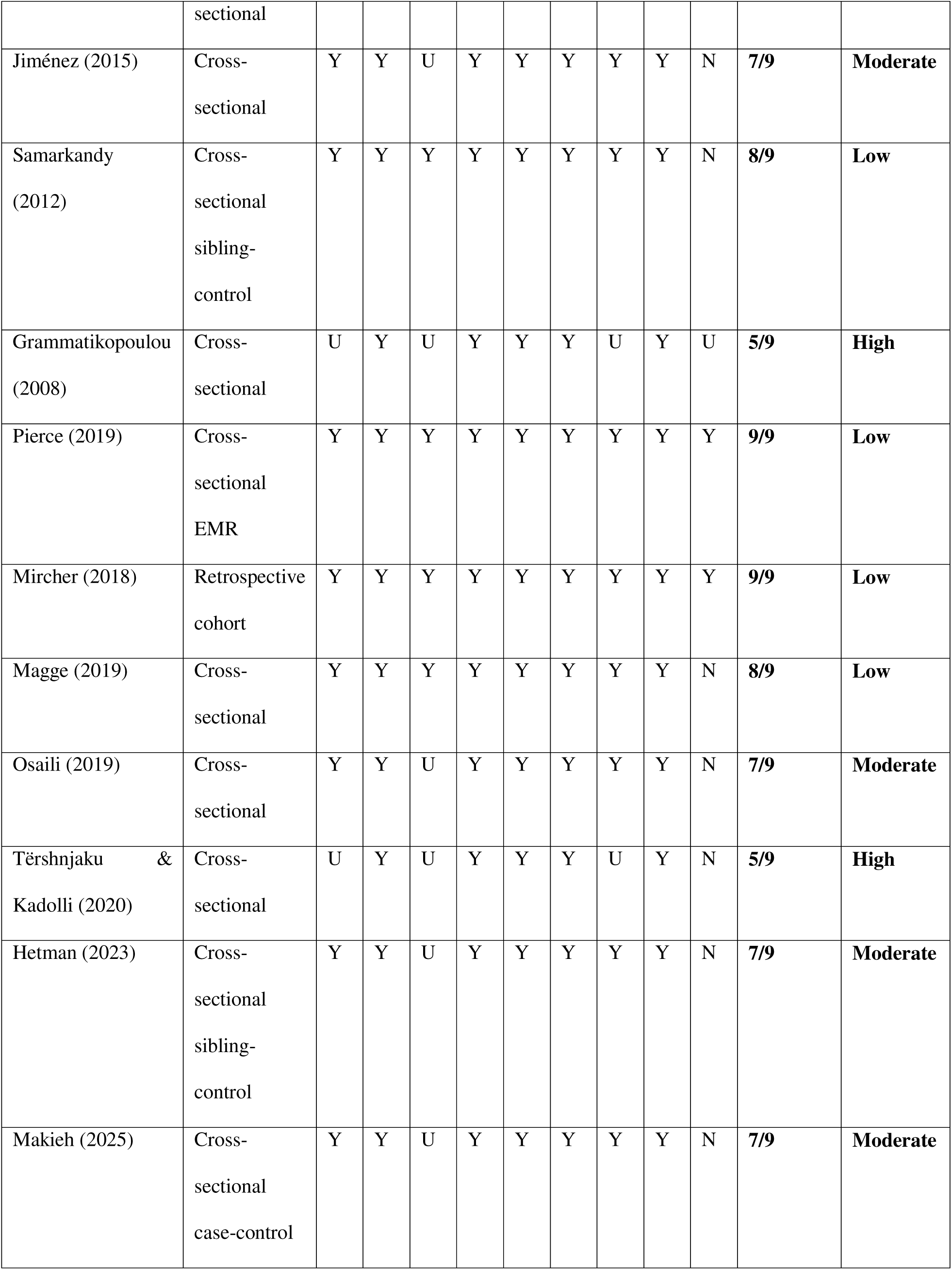
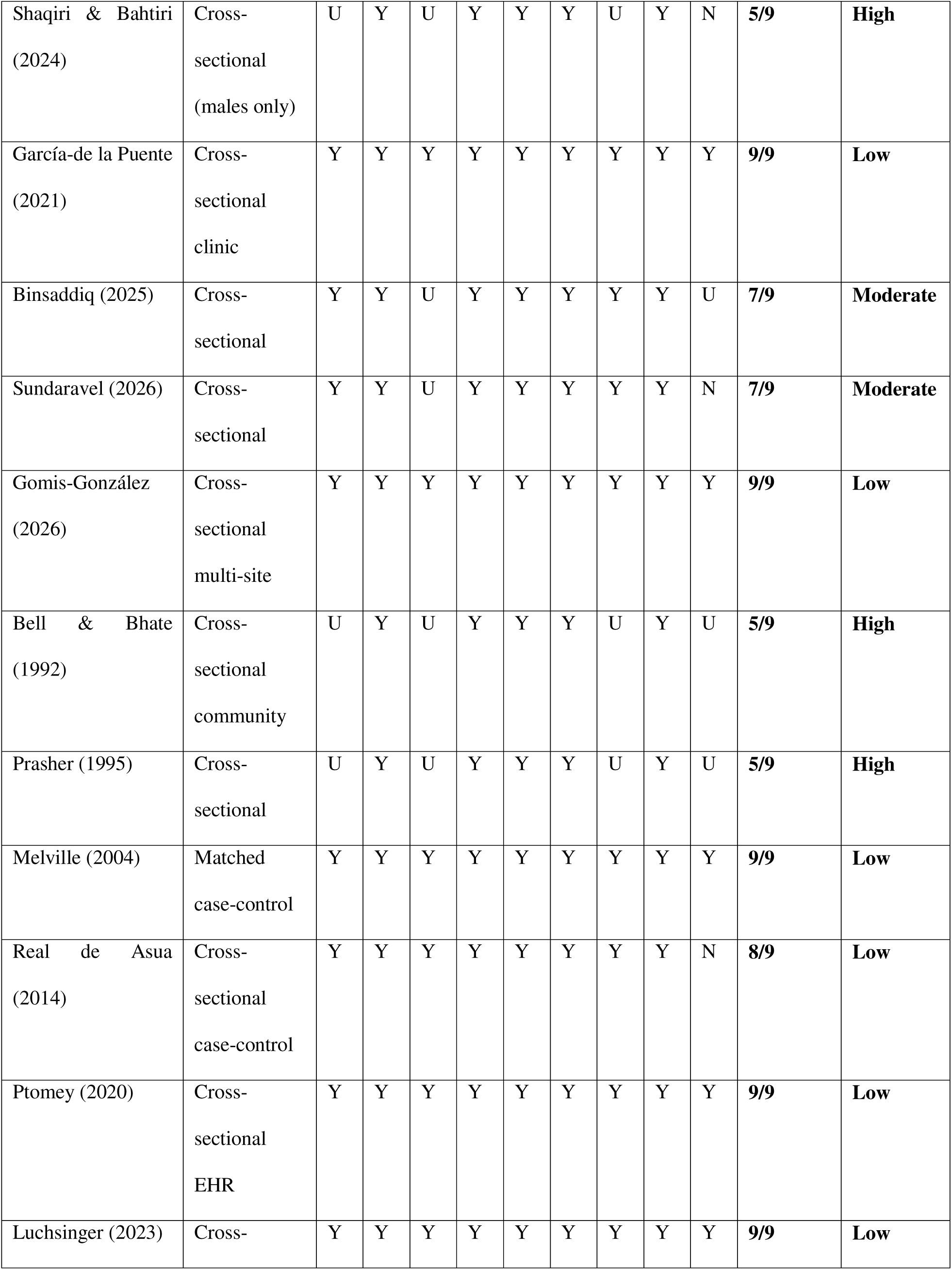
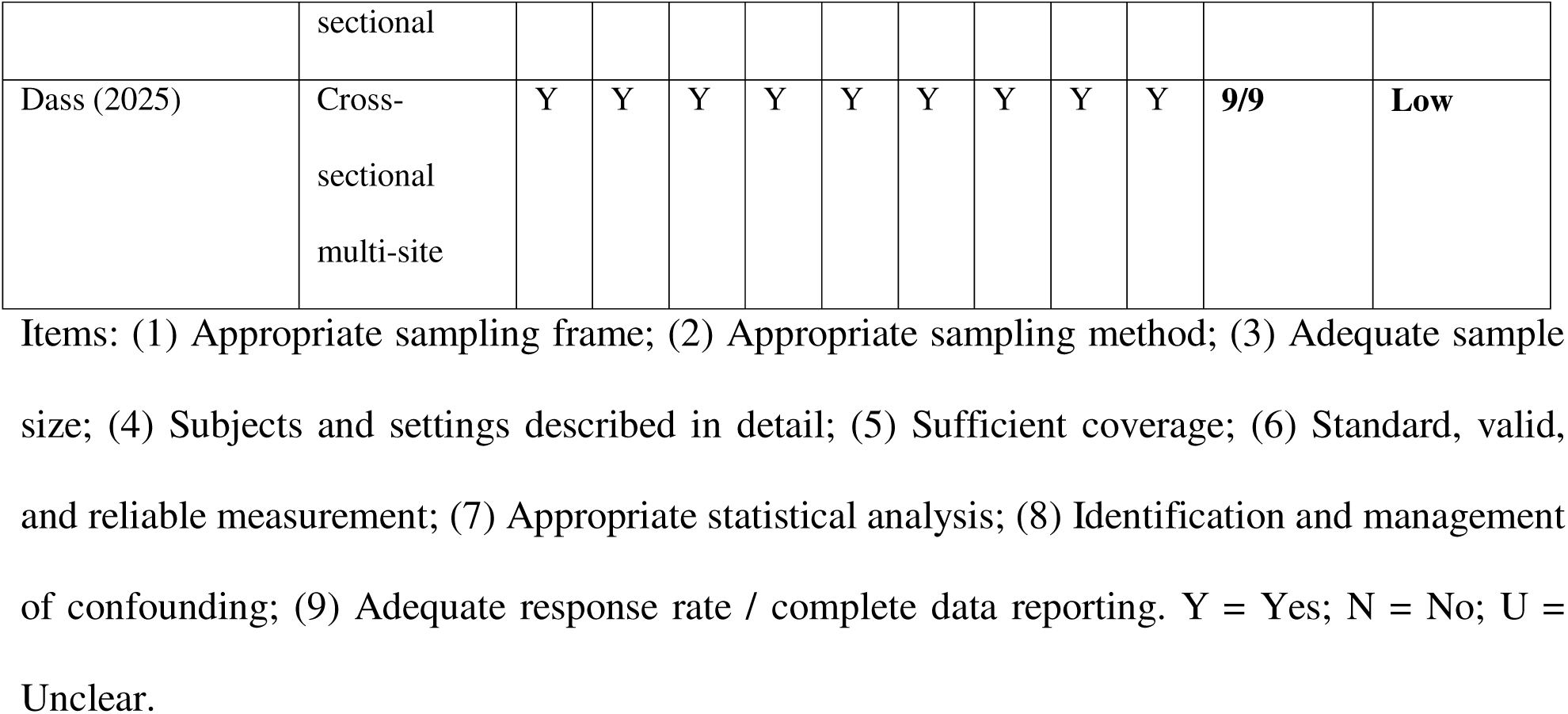
Risk of bias assessment using the JBI Critical Appraisal Checklist for Prevalence Studies (n = 26 studies)

### Certainty of Evidence

GRADE ratings are presented in Table 3. Certainty was rated Moderate for the primary prevalence outcomes and for the age-band subgroup analysis, downgraded one level from High due to serious inconsistency (I² = 88–95.3%) attributable principally to diagnostic criterion heterogeneity, age composition, and geographic setting. No further downgrading was warranted for risk of bias (majority of studies were low to moderate quality), indirectness (populations studied directly match the populations of interest), imprecision (combined N > 7,000 with narrow confidence intervals), or publication bias (Egger’s p = 0.6964). Certainty was rated Low for determinant associations and Very Low for the two-study OSA and NAFLD outcomes.

**Table 3.** GRADE summary of findings: certainty of the body of evidence for primary and secondary outcomes.

| Outcome | Studies (k/N) | Risk of Bias | Inconsistency | Indirectness | Imprecision | Publication Bias | Pooled Estimate | GRADE Certainty | Imp |
| --- | --- | --- | --- | --- | --- | --- | --- | --- | --- |
| Combined OW+OB prevalence — children/adolescents | 18 studies; N = 4,571 | Moderate concern (4 high-risk studies excluded in sensitivity analysis) | Serious ↓ ($I^2 = 88-93\%$; substantial heterogeneity) | Not serious | Not serious — $N > 4,000$ | Not serious — Egger $p = 0.6964$ | ~40–50% | MODER ATE $\oplus\oplus\oplus\boxminus$ | Crit |
| Age-band subgroup: children vs adults vs mixed | Forest plot analysis; N = 10,722 | Not serious | Serious ↓ ($I^2 = 95.3\%$ overall); subgroup $I^2$: 93.6%/67.8%/94.2% | Not serious | Not serious | Not serious — Egger $p = 0.6964$ | Children 0.18; Adults 0.36; Mixed 0.30 | MODER ATE $\oplus\oplus\oplus\boxminus$ | Crit |
| Obesity-only prevalence — adults | 8 studies; N = 1,682 | Not serious | Serious ↓ ($I^2 > 80\%$) | Not serious | Not serious | Not serious | ~48–75% | MODER ATE $\oplus\oplus\oplus\boxminus$ | Crit |
| Determinants of excess adiposity | ≥5 studies (multivariate analysis) | Moderate concern | Serious ↓ | Not serious | Serious ↓ (few studies) | Not assessed ($k < 10$) | Varied by factor | LOW $\oplus\oplus\boxminus\boxminus$ | Imp |
| Association: obesity and OSA / NAFLD in DS | 2 studies | Not serious | Not assessed ($k < 10$) | Not serious | Serious ↓ ($k = 2$) | Not assessed | RR 2.4 OSA; NAFLD 82% vs 45% | VERY LOW $\oplus\boxminus\boxminus\boxminus$ | Imp |

DS = Down syndrome; NAFLD = non-alcoholic fatty liver disease; OB = obesity; OSA =
obstructive sleep apnoea; OW = overweight. ↓ = downgraded by one level. GRADE certainty
levels: High $\oplus\oplus\oplus\oplus$ ; Moderate $\oplus\oplus\oplus\boxminus$ ; Low $\oplus\oplus\boxminus\boxminus$ ; Very Low $\oplus\boxminus\boxminus\boxminus$ .

## Discussion

### Principal findings

This systematic review and meta-analysis generates the first formally pooled global prevalence estimates of overweight and obesity in people with DS and identifies age band and diagnostic criterion as the two principal sources of heterogeneity across the included literature. The pooled prevalence of 18% in children and adolescents rising to 36% in adults, with a statistically significant subgroup difference (χ² = 14.39, p = 0.0007), indicates that the obesity burden in DS increases substantially across the lifespan and substantially exceeds general-population prevalence at every age stratum. Three studies that applied both a DS-specific growth chart and a general-population reference to the same cohorts found differences of 14 to 37% points in reported combined overweight and obesity prevalence [3–5], demonstrating that a substantial proportion of the heterogeneity previously attributed to biological or geographic variation across studies is attributable to inconsistent reference chart selection. These two findings have direct implications for how prevalence data in DS are collected, reported, and interpreted.

### The lifespan trajectory of obesity in DS

The age-progressive nature of excess adiposity in DS reflects the compounding of biological and environmental determinants that operate differentially across life stages. In the paediatric period, excess adiposity in DS has a well-characterised biological substrate. Leptin resistance, documented in prepubertal children with DS at adiposity levels comparable to unaffected siblings, indicates that central satiety signalling is impaired independently of accumulated fat mass [7]. A lower resting metabolic rate, arising from hypothalamic dysregulation, thyroid axis dysfunction, and reduced skeletal muscle mass secondary to hypotonia, establishes a structural metabolic disadvantage that operates throughout life [6]. These mechanisms are consistent with the observation from a sibling case-control study in Riyadh that overweight and obesity rates were markedly higher in children with DS than in matched siblings despite no significant difference in total energy intake or screen time between groups [24], confirming that reduced energy expenditure rather than excess intake is the dominant driver of early adiposity in this population.

In adulthood, additional factors compound the biological substrate established in childhood. Hypothyroidism, whose prevalence increases with age in DS, independently suppresses metabolic rate and promotes adipogenesis [2]. Progressive sarcopenia secondary to hypotonia and reduced habitual activity reduces resting energy expenditure further. Structured physical activity, which may be partially maintained through school-based programmes, typically diminishes after educational transitions without equivalent replacement in adult care pathways. The GO-DS21 multi-country cohort, enrolling 185 individuals aged 12 to 45 years across three European countries, found that age was the strongest independent predictor of BMI category after adjusting for sex, country, physical activity, and parental BMI, with each additional year of age associated with incrementally higher adiposity [9]. This finding is consistent with the Leicestershire Learning Disabilities Register data showing that obesity prevalence in adults with DS in the fourth and fifth decades of life substantially exceeded that in younger adults from the same register [36]. Taken together, these data indicate that the obesity trajectory in DS is progressive across the full adult lifespan and that adult health supervision frameworks require weight management components of equivalent specificity to those recommended for paediatric populations.

### Diagnostic criterion validity and the growth reference problem

The 14 to 37 percentage-point difference in prevalence generated by applying DS-specific versus general-population references to the same cohorts reflects a fundamental validity limitation of currently available DS-specific growth charts. The Myrelid SDM/2002, Cronk 1988, and Zemel/AAP 2015 charts were each constructed from reference populations that themselves carry excess adiposity, such that the normative distribution they encode does not represent a healthy weight standard but rather the weight distribution of an already-overweight reference group. Classification against such a reference measures relative standing within an elevated distribution rather than cardiometabolic risk, and therefore systematically misclassifies children with clinically significant adiposity as normal weight.

The clinical consequences of this misclassification are evident from the NAFLD data reported by Valentini and colleagues[11], in which 45% of children with DS classified as non-obese had hepatic steatosis detectable by ultrasound, compared with 5.7% of non-obese European children without DS. Children classified as weight-normal by DS-specific chart criteria were already accumulating hepatic steatosis at nine times the background rate of the general non-obese paediatric population. Until a DS-specific growth reference grounded in a non-overweight sample and validated against cardiometabolic endpoints is developed, general-population BMI references provide the only available instrument that indexes metabolic risk rather than relative adiposity within an elevated reference distribution. Standardised use of general-population references, with explicit reporting of the criterion applied in every publication, would substantially reduce the diagnostic heterogeneity that currently limits comparability across the DS obesity literature.

### Cardiometabolic consequences of excess adiposity

OSA in DS has an anatomical predisposition independent of weight, arising from midface hypoplasia, macroglossia, and pharyngeal hypotonia. Obesity amplifies OSA severity through pharyngeal fat deposition, as evidenced by the greater than twofold elevation in OSA risk associated with obesity in the Cincinnati retrospective cohort (RR 2.4; 95% CI 1.34 to 4.34), where obese children with DS were additionally more likely to have moderate or severe rather than mild OSA [10]. Given the anatomical predisposition that exists independent of weight, OSA surveillance in children with DS is warranted across all BMI categories, and obesity should be addressed as a modifiable amplifier of OSA severity within integrated management pathways rather than as a separate clinical concern.

The NAFLD findings warrant separate consideration because the mechanism appears partly independent of adiposity. In the Italian cohort, steatosis severity correlated positively with BMI (r = 0.37), waist circumference (r = 0.37), leptin (r = 0.35), and LDL-cholesterol (r = 0.22), and negatively with adiponectin (r = −0.23), consistent with an adipokine-mediated pathway (Valentini et al., 2017). TNF-α levels in non-obese children with DS in the same cohort exceeded published normative values for non-obese children without DS, suggesting that chromosome 21 gene dosage effects on inflammatory signalling contribute to hepatic steatosis independently of adiposity. These observations collectively support extending NAFLD surveillance in DS beyond those who are overweight or obese, with biochemical and imaging assessment beginning in middle childhood.

Adults with DS have high rates of dyslipidaemia, insulin resistance, and obesity, yet overt atherosclerotic cardiovascular events are less common than would be predicted from their risk factor burden, a pattern attributed to chromosome 21 gene dosage effects on lipoprotein oxidation, chronically lower ambient blood pressure, and minimal lifetime tobacco exposure. This relative protection from atherosclerotic events does not, though, extend to OSA-mediated pulmonary hypertension, NAFLD-related hepatic fibrosis, musculoskeletal disability secondary to sarcopenic obesity, or the cognitive consequences of excess adiposity. Cross-sectional data from the Alzheimer’s Biomarker Consortium-Down Syndrome study demonstrated that obesity and physical inactivity independently impaired cognitive performance in adults with DS, and that physically active obese adults scored better on episodic memory, executive function, and dementia symptom measures than sedentary non-obese adults [14]. These findings indicate that physical activity modifies the cognitive impact of adiposity in DS and that both factors require concurrent management in adult care pathways.

### Determinants of excess adiposity and intervention implications

The consistent finding across multiple studies that total caloric intake does not differ significantly between overweight and normal-weight individuals with DS identifies reduced energy expenditure rather than hyperphagia as the primary modifiable metabolic target. Physical inactivity is the most consistently documented behavioural determinant of higher adiposity in DS across all age groups. Accelerometry combined with DXA in 77 youth with DS found that light physical activity, rather than MVPA, was most strongly associated with lower visceral fat accumulation, and that youth with DS accumulate less MVPA than typically developing peers [8]. In the GO-DS21 cohort, lower physical activity was a significant independent predictor of higher BMI across three countries while total caloric intake did not differ across BMI categories, and protein intake was the only macronutrient inversely associated with overweight [9]. A prospective two-year study in 44 children with DS in Verona found that structured dietary and physical activity guidance delivered by a paediatrician produced significant reductions in BMI z-scores and improved activity scores, though lipid profiles required separate management [42]. A meta-analysis of 15 randomised controlled trials confirmed that aerobic training reduces fat mass (SMD = −0.44) and waist circumference (SMD = −0.39) in people with DS, with changes in body composition more pronounced than changes in BMI, supporting body composition rather than weight alone as the primary monitoring outcome in intervention research [41].

Parental BMI was a significant independent predictor of child BMI in the GO-DS21 analysis [9], identifying household-level behavioural determinants that operate beyond DS-specific physiology. Family-based intervention designs that include parental dietary and activity components are likely to address these determinants more effectively than programmes targeting only the person with DS. No pharmacological weight management trial has been conducted in DS populations, representing a gap in the evidence base that is proportionally large given the scale of the burden documented in this review.

### Geographic evidence gaps

The absence of eligible primary prevalence data from sub-Saharan Africa across three independent systematic search strategies identifies a critical gap in the global evidence base. Sub-Saharan Africa accounts for a substantial proportion of global DS births, a share amplified by limited prenatal screening access [1], yet the nutritional and cardiometabolic profile of people with DS in this region is entirely unknown from the published literature. The lower prevalence estimates from middle-income settings, including 15.3% from a specialist DS clinic in Mexico City [31] and 31.6% from a tertiary referral hospital in New Delhi [33], suggest that active nutritional surveillance through dedicated DS specialist pathways attenuates the adiposity burden relative to the higher estimates from community and electronic health record cohorts in high-income countries. Investment in primary epidemiological research and DS specialist infrastructure in sub-Saharan Africa and South Asia would simultaneously generate the evidence base currently absent from this literature and deliver the clinical monitoring most likely to reduce prevalence in those settings.

### Strengths and limitations

This review searched six databases without date or language restriction, enrolled both paediatric and adult populations, and applied a pre-specified age-band subgroup analysis that moves beyond the unannotated 23 to 70% range cited in the 2016 narrative review by Bertapelli and colleagues. GRADE certainty ratings are provided for all primary and secondary outcomes.

Between-study heterogeneity was high (I² = 95.3%; prediction interval 7 to 53%), as expected given the diversity of diagnostic criteria, age compositions, settings, and data collection periods across included studies. The age-band subgroup and diagnostic criterion family together explain a meaningful portion of this variance, but substantial residual heterogeneity within each stratum indicates that additional sources including institutional setting, comorbidity composition, and data collection era also contribute. Five high-risk studies were retained in the primary analysis; their exclusion in sensitivity analyses did not materially alter any pooled estimate. Individual patient data were unavailable, preventing within-study sex-stratified and age-stratified analyses. The OSA and NAFLD associations each rest on a single contributing study, precluding pooled effect estimates and limiting causal inference.

### Conclusions

Overweight and obesity in DS are highly prevalent, worsen substantially with age, and exceed general-population rates at every life stage. Roughly one in five people with DS is affected overall and more than one in three adults. The reference chart applied to the same cohort changes the reported prevalence by up to 37 percentage points; DS-specific growth charts derived from overweight reference populations should not serve as the primary classification standard until a cardiometabolic-validated alternative is developed. OSA and NAFLD occur at elevated rates in DS regardless of weight status and warrant surveillance based on diagnosis rather than BMI category. Physical activity promotion, protein-adequate dietary support, and family-based behavioural programmes are the most evidence-supported intervention targets. The absence of primary prevalence data from sub-Saharan Africa is the most consequential geographic gap in this literature. DS health supervision guidelines should specify quantitative overweight thresholds, age-stratified intervention triggers, and structured dietitian referral criteria as the most immediate clinical policy actions arising from this synthesis.

## Data Availability

All data produced in the present work are contained in the manuscript

## Supporting Information

S1 File. PRISMA 2020 checklist with page-number citations.

S2 File. Full database-specific search strings.

## Notes

### Competing Interest Statement

The authors have declared no competing interest.

