## Supplementary material for "Prevalence, determinants, and cardiometabolic consequences of overweight and obesity among people with Down syndrome: a systematic review and meta-analysis": S1 File

**Full Database Search Strings**

### 1. PubMed/MEDLINE

| **No** | **Search Strategy** | **Description** |
| --- | --- | --- |
| #1 | ("Down Syndrome"[Mesh] OR "Trisomy 21"[Mesh] OR "down syndrome"[tiab] OR "trisomy 21"[tiab] OR "trisomy-21"[tiab] OR "DS"[tiab] OR "trisomy21"[tiab]) | Down syndrome / trisomy 21 |
| #2 | ("Overweight"[Mesh] OR "Obesity"[Mesh] OR "Pediatric Obesity"[Mesh] OR "Body Mass Index"[Mesh] OR "Body Composition"[Mesh] OR "Adipose Tissue"[Mesh] OR overweight[tiab] OR over-weight[tiab] OR obes*[tiab] OR adipos*[tiab] OR "body mass index"[tiab] OR BMI[tiab] OR "body fat"[tiab] OR "fat mass"[tiab] OR "waist circumference"[tiab] OR "waist-to-height"[tiab] OR skinfold*[tiab] OR "bioelectrical impedance"[tiab] OR "bio-electrical impedance"[tiab] OR "body composition"[tiab] OR "dual energy x-ray"[tiab] OR "dual-energy x-ray"[tiab] OR DEXA[tiab] OR DXA[tiab] OR "weight gain"[tiab] OR "weight status"[tiab] OR "weight-for-height"[tiab] OR "body weight"[tiab] OR "growth chart"[tiab] OR "growth charts"[tiab]) | Overweight, obesity, and adiposity-related terms |
| #3 | ("Prevalence"[Mesh] OR "Epidemiology"[Mesh] OR "Cross-Sectional Studies"[Mesh] OR "Cohort Studies"[Mesh] OR "Case-Control Studies"[Mesh] OR prevalence[tiab] OR incidence[tiab] OR frequency[tiab] OR proportion[tiab] OR cross-sectional[tiab] OR "cross sectional"[tiab] OR cohort[tiab] OR "case-control"[tiab] OR "case control"[tiab] OR observational[tiab] OR survey[tiab] OR epidemiolog*[tiab]) | Observational study designs and prevalence terms |
| #4 | #1 AND #2 | Down syndrome + adiposity |
| #5 | #4 AND #3 | Final: Down syndrome + adiposity + observational design |

### 2. EMBASE (via Elsevier)

| **No** | **Search Strategy** | **Description** |
| --- | --- | --- |
| #1 | ('down syndrome'/exp OR 'trisomy 21'/exp OR 'down syndrome':ab,ti OR 'trisomy 21':ab,ti OR 'trisomy-21':ab,ti OR 'DS':ab,ti) | Down syndrome / trisomy 21 |
| #2 | ('overweight'/exp OR 'obesity'/exp OR 'body mass index'/exp OR 'body composition'/exp OR 'adipose tissue'/exp OR overweight:ab,ti OR obes*:ab,ti OR adipos*:ab,ti OR 'body mass index':ab,ti OR BMI:ab,ti OR 'body fat':ab,ti OR 'fat mass':ab,ti OR 'waist circumference':ab,ti OR skinfold*:ab,ti OR 'bioelectrical impedance':ab,ti OR 'body composition':ab,ti OR DXA:ab,ti OR DEXA:ab,ti OR 'weight gain':ab,ti OR 'growth chart*':ab,ti) | Overweight, obesity, and adiposity-related terms |
| #3 | ('prevalence'/exp OR 'cross-sectional study'/exp OR 'cohort analysis'/exp OR 'case control study'/exp OR prevalence:ab,ti OR 'cross-sectional':ab,ti OR cohort:ab,ti OR 'case-control':ab,ti OR observational:ab,ti OR survey:ab,ti OR epidemiolog*:ab,ti) | Observational study designs and prevalence terms |
| #4 | #1 AND #2 | Down syndrome + adiposity |
| #5 | #4 AND #3 | Final combined set |

### 3. Scopus

| **No** | **Search Strategy** | **Description** |
| --- | --- | --- |
| #1 | (TITLE-ABS-KEY("down syndrome" OR "trisomy 21" OR "trisomy-21")) | Down syndrome / trisomy 21 |
| #2 | (TITLE-ABS-KEY(overweight OR obes* OR adipos* OR "body mass index" OR BMI OR "body fat" OR "fat mass" OR "waist circumference" OR skinfold* OR "bioelectrical impedance" OR "body composition" OR DXA OR DEXA OR "weight gain" OR "growth chart")) | Overweight, obesity, and adiposity-related terms |
| #3 | (TITLE-ABS-KEY(prevalence OR "cross-sectional" OR cohort OR "case-control" OR observational OR survey OR epidemiolog*)) | Observational study designs |
| #4 | #1 AND #2 | Down syndrome + adiposity |
| #5 | #4 AND #3 | Final combined set |

### 4. Web of Science Core Collection

| **No** | **Search Strategy** | **Description** |
| --- | --- | --- |
| #1 | TS=("down syndrome" OR "trisomy 21" OR "trisomy-21" OR "DS") | Down syndrome / trisomy 21 |
| #2 | TS=(overweight OR obes* OR adipos* OR "body mass index" OR "BMI" OR "body fat" OR "fat mass" OR "waist circumference" OR "skinfold" OR "bioelectrical impedance" OR "body composition" OR "DXA" OR "DEXA" OR "weight gain" OR "growth chart") | Overweight, obesity, and adiposity-related terms |
| #3 | TS=(prevalence OR "cross-sectional" OR cohort OR "case-control" OR observational OR survey OR epidemiolog*) | Observational designs |
| #4 | #1 AND #2 | Down syndrome + adiposity |
| #5 | #4 AND #3 | Final combined set |

### 5. CINAHL (Cumulative Index to Nursing and Allied Health Literature)

| **No** | **Search Strategy** | **Description** |
| --- | --- | --- |
| S1 | (MH "Down Syndrome") OR TI ( "down syndrome" OR "trisomy 21" ) OR AB ( "down syndrome" OR "trisomy 21" ) | Down syndrome / trisomy 21 |
| S2 | (MH "Obesity") OR (MH "Overweight") OR (MH "Body Mass Index") OR (MH "Body Composition") OR TI ( overweight OR obes* OR adipos* OR "body mass index" OR BMI OR "body fat" OR "waist circumference" OR skinfold* OR "bioelectrical impedance" OR "body composition" OR DXA OR DEXA OR "weight gain" OR "growth chart" ) OR AB ( overweight OR obes* OR adipos* OR "body mass index" OR BMI OR "body fat" OR "waist circumference" OR skinfold* OR "bioelectrical impedance" OR "body composition" OR DXA OR DEXA OR "weight gain" OR "growth chart" ) | Overweight, obesity, and adiposity-related terms |
| S3 | (MH "Cross Sectional Studies") OR (MH "Cohort Studies") OR TI ( prevalence OR "cross-sectional" OR cohort OR "case-control" OR observational OR survey OR epidemiolog* ) OR AB ( prevalence OR "cross-sectional" OR cohort OR "case-control" OR observational OR survey OR epidemiolog* ) | Observational study designs |
| S4 | S1 AND S2 | Down syndrome + adiposity |
| S5 | S4 AND S3 | Final combined set |

### 6. LILACS (Latin American and Caribbean Health Sciences Literature)

| **No** | **Search Strategy** | **Description** |
| --- | --- | --- |
| #1 | ("down syndrome" OR "sindrome de down" OR "síndrome de down" OR "trisomy 21" OR "trisomia 21" OR "trissomia do 21") [Words] | Down syndrome / trisomy 21 (English, Spanish, Portuguese) |
| #2 | (overweight OR "sobrepeso" OR obes* OR adipos* OR "body mass index" OR "indice de masa corporal" OR "índice de massa corporal" OR BMI OR "body fat" OR "waist circumference" OR "circunferencia de cintura" OR "bioelectrical impedance" OR "body composition" OR DXA OR "growth chart" OR "curva de crescimento") [Words] | Overweight, obesity, and adiposity-related terms (English, Spanish, Portuguese) |
| #3 | (prevalence OR prevalencia OR prevalência OR "cross-sectional" OR "corte transversal" OR cohort OR cohorte OR coorte OR "case-control" OR observational OR epidemiolog*) [Words] | Observational designs (English, Spanish, Portuguese) |
| #4 | #1 AND #2 | Down syndrome + adiposity |
| #5 | #4 AND #3 | Final combined set |
