## Supplementary material for "Prevalence, determinants, and cardiometabolic consequences of overweight and obesity among people with Down syndrome: a systematic review and meta-analysis": S2 File

**PRISMA-P 2015 Checklist**

| **Section and Topic** | **Item No** | **Checklist Item** | **Location in Protocol** |
| --- | --- | --- | --- |
| **ADMINISTRATIVE INFORMATION** | | | |
| **Title** | | | |
|  | **1a** | Identify the report as a protocol of a systematic review | Title page — title contains "protocol for a systematic review and meta-analysis" |
|  | **1b** | If the protocol is for an update of a previous systematic review, identify as such | N/A — this is a new protocol with no prior version |
| **Registration** | | | |
|  | **2** | If registered, provide the name of the registry (e.g., PROSPERO) and registration number | Abstract — PROSPERO registration pending; number to be inserted prior to publication |
| **Authors** | | | |
|  | **3a** | Provide name, institutional affiliation, e-mail address of all protocol authors; provide physical mailing address of corresponding author | Title page — Ivaan Pitua (Makerere University;; ORCID: 0000-0003-0768-1315); Felix Bongomin (Gulu University) |
|  | **3b** | Describe contributions of protocol authors and identify the guarantor of the review | Title page — IP is the guarantor; both authors contributed to protocol design, search strategy development, and manuscript drafting |
| **Amendments** | | | |
|  | **4** | If the protocol represents an amendment of a previously completed or published protocol, identify as such; otherwise, state plan for documenting important protocol amendments | N/A — original protocol; substantive amendments will be documented with date and rationale in the PROSPERO record |
| **Support** | | | |
|  | **5a** | Indicate sources of financial or other support for the review | Methods — no external funding; no conflict of interest declared |
|  | **5b** | Provide name for the review funder and/or sponsor | N/A — unfunded |
|  | **5c** | Describe roles of funder(s), sponsor(s), and/or institution(s), if any, in developing the protocol | N/A — unfunded |
| **INTRODUCTION** | | | |
|  | **6** | Describe the rationale for the review in the context of what is already known | Introduction — paragraphs 1–5 establish the epidemiological burden of overweight and obesity in DS, identify the methodological heterogeneity of existing estimates, and define the evidence gap (no prior pooled global estimate) |
|  | **7** | Provide an explicit statement of the question(s) the review will address with reference to participants, interventions, comparators, and outcomes (PICO) | Introduction, final paragraph — PICO: People with Down syndrome (any age) \| Overweight or obesity (any diagnostic criterion) \| General population or non-obese DS comparator \| Pooled prevalence; determinants; adverse health outcomes |
| **METHODS** | | | |
| **Eligibility Criteria** | | | |
|  | **8** | Specify the study characteristics (such as PICO, study design, setting, time frame) and report characteristics (such as years considered, language, publication status) to be used as criteria for eligibility for the review | Methods > Eligibility Criteria — includes observational designs (cross-sectional, cohort, case-control, RCT baselines); any setting; any country; any age group; no date or language restriction; n ≥ 10 DS participants with extractable prevalence data required |
| **Information Sources** | | | |
|  | **9** | Describe all intended information sources (such as electronic databases, contact with study authors, trial registers, or other grey literature sources) with planned dates of coverage | Methods > Information Sources — PubMed/MEDLINE, EMBASE, Scopus, Web of Science, CINAHL, LILACS; Google Scholar (first 200 results); reference mining of included studies and key prior reviews; no date restriction; search date: May 2026 |
| **Search Strategy** | | | |
|  | **10** | Present draft of search strategy to be used for at least one electronic database, including planned limits, such that it could be repeated | Methods > Search Strategy (PubMed example string) and S1 File (full strings for all 6 databases) |
| **Study Records** | | | |
|  | **11a** | Describe the mechanism(s) that will be used to manage records and data throughout the review | Methods > Study Selection — Covidence systematic review software for deduplication, screening, and record management |
|  | **11b** | State the process that will be used for selecting studies through each phase of the review (screening, eligibility, inclusion in meta-analysis) | Methods > Study Selection — two independent reviewers screen titles/abstracts then full texts; disagreements resolved by discussion or third-reviewer adjudication; PRISMA 2020 flow diagram will document all stages |
|  | **11c** | Describe planned method of extracting data from reports, any processes for obtaining and confirming data from investigators | Methods > Data Extraction — standardised Microsoft Excel form, piloted on five studies; two independent reviewers; corresponding authors contacted for missing data |
| **Data Items** | | | |
|  | **12** | List and define all variables for which data will be sought, including any pre-planned data assumptions and simplifications | Methods > Data Extraction — variables listed: author/year, country, income tier, study design, data collection period, setting, n, age, sex, karyotype confirmation, diagnostic criterion, measurement method, OW prevalence, OB prevalence, combined OW+OB prevalence, comorbidity profile, determinant data, secondary outcome data |
| **Outcomes and Prioritisation** | | | |
|  | **13** | List and define all outcomes for which data will be sought, including prioritisation of main and additional outcomes, with rationale | Methods > Eligibility Criteria > Outcomes — Primary: pooled prevalence of overweight, obesity, and combined OW+OB. Secondary: determinants of excess adiposity; associations with obstructive sleep apnoea, NAFLD, dyslipidaemia, insulin resistance, all-cause mortality |
| **Risk of Bias** | | | |
|  | **14** | Describe anticipated methods for assessing risk of bias of individual studies, including whether this will be done at the outcome or study level, or both; state how this information will be used in data synthesis | Methods > Quality Assessment — JBI Prevalence Studies checklist (primary outcome); Newcastle-Ottawa Scale (secondary outcome associations); assessed at study level; incorporated in sensitivity analyses and GRADE certainty assessment; no studies excluded solely on quality |
| **Data Synthesis** | | | |
|  | **15a** | Describe criteria under which study data will be quantitatively synthesised | Methods > Data Synthesis > Prevalence Analysis — quantitative synthesis performed if ≥ 2 studies share comparable population and outcome definition; random-effects model used throughout |
|  | **15b** | If data are appropriate for quantitative synthesis, describe planned summary measures, methods of handling data, and methods of combining data from studies, including any planned exploration of consistency | Methods > Data Synthesis > Prevalence Analysis — Freeman-Tukey double-arcsine transformation; DerSimonian-Laird random-effects model; I² and τ² for heterogeneity; subgroup analyses by age, criterion, region, income tier, sex, study period |
|  | **15c** | Describe any proposed additional analyses (such as sensitivity or subgroup analyses, meta-regression) | Methods > Data Synthesis > Subgroup Analyses, Meta-regression, Sensitivity Analyses — pre-specified subgroup by age band, criterion family, region, income tier, sex, study decade; meta-regression with mean age, study year, % female, % with hypothyroidism (≥10 studies per covariate); sensitivity by risk-of-bias category, age group, criterion type, sample size, and leave-one-out |
|  | **15d** | If quantitative synthesis is not appropriate, describe the type of summary planned | N/A — quantitative synthesis is anticipated to be appropriate; narrative synthesis applied as fallback if <2 studies per stratum |
| **Meta-bias** | | | |
|  | **16** | Specify any planned assessment of meta-bias(es) (such as publication bias across studies, selective reporting within studies) | Methods > Publication Bias — funnel plot visual inspection and Egger's regression test if ≥ 10 studies per analysis; trim-and-fill method if asymmetry detected; limitations of these tests for proportion meta-analyses acknowledged |
| **Confidence in Cumulative Evidence** | | | |
|  | **17** | Describe how the strength of the body of evidence will be assessed (such as GRADE) | Methods > Assessment of Certainty of Evidence — GRADE approach for secondary outcome associations (two independent reviewers; five GRADE domains; GRADEpro GDT for Summary of Findings tables); JBI appraisal ratings summarised for primary prevalence estimates |

** It is strongly recommended that this checklist be read in conjunction with the PRISMA-P 2015 Explanation and Elaboration document. Source: Shamseer L, Moher D, Clarke M, et al. Preferred reporting items for systematic review and meta-analysis protocols (PRISMA-P) 2015: elaboration and explanation. BMJ. 2015;349:g7647.*
